# Acceptability and feasibility of maternal screening for Group B Streptococcus: a rapid review

**DOI:** 10.1101/2024.06.28.24309381

**Authors:** Georgina Constantinou, Rebecca Webb, Susan Ayers, Eleanor J Mitchell, Jane Daniels

## Abstract

**Background:** The risks and benefits of maternal screening for GBS during pregnancy or the intrapartum period are widely debated, since screen positive results trigger prophylactic antibiotic use. There is little known about women’s and health professional’s views regarding GBS screening.

**Objectives:** To conduct a rapid review to synthesise evidence on women and health professionals’: (1) knowledge and awareness of; (2) preferences for; and (3) acceptability of GBS screening programmes, and (4) how feasible they are to implement.

**Method:** Literature searches were conducted using online databases from their inception to 2023. Papers were included if they reported primary research from the perspectives of health professionals and women, about their knowledge and awareness, preferences, acceptability and feasibility of different types of GBS screening programmes. Data were assessed for confidence using GRADE-CERQual and analysed using a convergent synthesis approach.

**Findings:** 42 papers were eligible for inclusion. A total of 16,306 women and professionals were included. Women generally did not have extensive knowledge about GBS. Health professionals had a higher level of knowledge than women. Women were generally (but not universally) positive about GBS testing procedures. Some women were concerned about the impact on their place of birth.

**Discussion and Conclusion:** Where GBS screening programmes are available, parents must be provided with high quality information about them. Health professionals and service managers need to weigh up the benefits and risks of screening for GBS with local feasibility and treatment options, and with women’s individual values and birth plans.

**Statement of significance:** **Problem:** Maternal GBS colonisation at birth can lead to invasive GBS disease. The risks and benefits of screening for GBS during pregnancy is widely debated.

**What is already known:** Different countries use different GBS screening strategies, such as the universal screening strategy vs risk based.

**What this paper adds:** The World Health Organization reviewed their GBS policy guidelines in 2024.

**Results** from this paper were used to ensure women and health professional’s views were considered. This paper found that women are generally (but not universally) positive about GBS testing procedures with some concerned about the impact on their birth choices.

## Introduction

Group B Streptococcus (GBS) is a bacteria that is typically harmless with no symptoms, although if passed to the baby during birth it can result in infection which can be fatal for the infant if untreated.^1, 2^ Globally it is estimated that around 1 in 5 pregnant women are carriers of GBS bacteria.^1^ Estimates show that there were 319,000 cases of invasive GBS disease globally; 205,000 were early onset GBS (EOGBS) and 114,000 cases of late onset GBS (LOGBS), with an estimation of 90,000 infants deaths worldwide.^1^

In many high-income maternity care settings, if maternal GBS colonisation is suspected or detected, maternal intrapartum antibiotic prophylaxis (IAP) is given during labour to reduce transmission to the baby in an effort to prevent EOGBS disease.^3^ However it is unclear if women should be targeted based on the presence of clinical risk factors or by screening for GBS colonisation during pregnancy,^3^ therefore internationally, detection strategies vary.^4^ Some countries deploy universal GBS screening approaches whereby all pregnant women are offered GBS testing in pregnancy such as the USA.^4^ Whereas other countries identify women with certain clinical risk factors (e.g., GBS bacteria in current or previous pregnancy, women with risk of pre-term birth, women with a temperature of 38°C or higher^5^) and then prescribe IAP such as the Netherlands^6^ and UK^7^ There is also combination strategies, whereby all women are screened, but only those with positive GBS results and a risk factor are offered IAP.^8^ Variation in approach is partly because women who screen positive during testing do not necessarily result in GBS being present at the time of birth, and vice versa. Missing potential GBS colonisation, and therefore failing to prevent neonatal illness or death, or widespread use of antibiotics for many women and neonates who are in fact GBS negative are debated concerns. Therefore, the ideal prevention strategy remains unclear and with rapid testing for GBS now available, this adds an additional element to these debates.^9^

No randomised controlled trials have previously looked at universal screening vs other types of screening programmes.^2, 10^ However, the GBS2 randomised trial tested the accuracy of the intrapartum test in diagnosing maternal GBS colonisation, compared to treatment as usual and found that the accuracy of the rapid test was acceptable^11^. Furthermore, the efficacy of implementing routine universal screening as opposed to a risk-factor based approach is currently being investigated in a large multi-centre clinical trial in the UK: the GBS3 trial.^12^

It is likely that results from these trials will be used to inform policy guidelines. For example, the World Health Organization reviewed their GBS policy guidelines in 2024, having last done so in 2015.^13^ It is important that women and healthcare professionals views are taken into account, as these are the people who will be impacted by changes to GBS policy and practice. This rapid review will therefore collate and synthesise the available evidence on women and healthcare professionals views on GBS screening strategies and provide a critical appraisal and overview of the evidence-base. These findings will be used to help inform the World Health Organizations development of GBS policy guidelines and can be used to help inform other organisations/services GBS policy and practice.

## Methods

### Aims

To conduct a rapid review to synthesise evidence on women and health professionals’: (1) knowledge and awareness of; (2) preferences for; and (3) acceptability of GBS screening programmes, and (4) how feasible they are to implement.

## Eligibility criteria

Studies were included if they reported primary research including perspectives of health professionals and women, knowledge and awareness of, preferences for, and acceptability of GBS screening programmes, as well as how feasible they are to implement. These variables were chosen as they are in line with the World Health Organization’s guidelines for developing guidelines.^14^ Both qualitative and quantitative studies were included. If quantitative they must have reported information on, knowledge, awareness, preferences, acceptability, feasibility/adherence of GBS screening to participants. Studies were excluded if they: (1) did not discuss GBS screening during pregnancy or birth; (2) were non-empirical papers; (3) were reviews.

## Information sources

The following online databases were searched from their inception to 2023: Academic Search Ultimate; Cumulative Index to Nursing and Allied Health Literature (CINAHL); EMBASE; Global Index Medicus; MEDLINE; PsychARTICLES; PsycINFO; PubMed; Scopus; Web of Science. The date of the last search was 12^th^ September 2023. Forward and backward searches of included studies were completed by the 5^th^ October 2023.

## Search

Searches were carried out using search terms that were combined with Boolean operators “OR” and “AND”. Search terms included, but were not limited to, women OR mother OR parent* AND pregnan* OR *natal OR *partum AND GBS OR Group B Strep OR GBS Bacteria AND test* OR screen* OR swab* AND value OR view OR experience (See supplementary material A for full search syntax).

## Review selection

Search results were imported into Eppi-Reviewer 4 and duplicates were removed by GC. The remaining papers were screened by title and abstract by GC. As per Cochrane Rapid Review guidelines,^15^ 20% (n = 257) of the title and abstracts were double screened by RW. Decisions to include or exclude were concordant in 69.6% of cases. Full text screening was carried out by RW, and as per Cochrane Rapid Review guidelines,^15^ 10 full texts were double screened by GC and RW. Decisions to include or exclude were concordant for 81.8% of cases. Furthermore, all excluded texts were assessed by GC to ensure they were not eligible for inclusion.

## Data collection process and data items

Each paper was read in full, and relevant parts of the text inputted into a Microsoft Excel spreadsheet by GC. The data that were extracted included the following: authors; year of publication; country; study design; sample size; participant characteristics; type of GBS testing; outcomes (knowledge, awareness, preferences, acceptability, feasibility/adherence); strengths and limitations. The researchers obtained or confirmed missing or ambiguous data by contacting authors.

## Critical appraisal of included studies

The quality of the included studies was assessed using: the questionnaire critical appraisal checklist^16^ for quantitative questionnaire papers; the Joanna Briggs Cross-Sectional Studies tool for other quantitative papers^17^; the Walsh and Downe^18^ tool for qualitative studies and; the Mixed Methods Appraisal Tool^19^ for mixed method papers. Although there is a move towards a domain based approach to critical appraisal^20^, the studies included used a variety of methodologies so domains for each critical appraisal tool would have been difficult to compare. It has been argued there is no evidence a checklist or domains-based approach is better than the other^21^, and that appraisal should be logically incorporated into the overall analysis.^22^ Therefore a score-based method was used allowing for comparisons across each study. Furthermore, no papers were excluded based on their methodological appraisal score, and in addition to using methodological appraisal ratings to determine confidence in individual papers, we also used the GRADE-CERQual approach^23^ to look at confidence in statement of findings identified from the review (see below). The use of multiple factors in determining the confidence of findings is recommended when rating the overall quality of a body of evidence^24^.

Therefore, each critical appraisal question for each paper was assigned a score of: 1 = Yes if the paper fully met the criteria; 0.5 = if the paper only partially met the criteria; and 0 = if the paper did not meet the criteria. A percentage was calculated by dividing the achieved score from the total possible score and multiplying by 100.

## Synthesis of results

Findings relevant to the study aims were extracted and recorded in Excel and summarised to show each of the included studies relevant findings. A convergent synthesis^25^ which allows for the mapping of the findings of studies from divergent methodological traditions and epistemological foundations into themes in relation to the aims of the review was used. For quantitative data means and standard deviations were calculated using Excel. There were not enough quantitative data to run analyses to account for the impact of location, national income level, screening programme, and health insurance/free health care on the outcomes in question. Themes were then refined further into statement of findings (see supplementary material B) and presented narratively.

## Assessment of confidence in the findings

The methodologies of papers 1-7 of the GRADE-CERQual series were used to evaluate confidence in the findings. Given the vast number of papers, and the different methodologies and aims used, certain rules were applied to allow for conclusions about confidence to be drawn (see Table 1). These rules are consistent with those used in a previous evidence synthesis regarding perinatal mental health care^26^. The group of papers underpinning each statement of findings was assessed on their methodological limitations^27^; coherence^28^, adequacy of data^29^, and relevance of data^30^ (see supplementary materials C-F). As per GRADE-CERQual the confidence of each of these four aspects was rated as: high confidence, moderate confidence, low confidence and very low confidence. A final evidence profile was developed (see supplementary materials B).

**Table 1.**
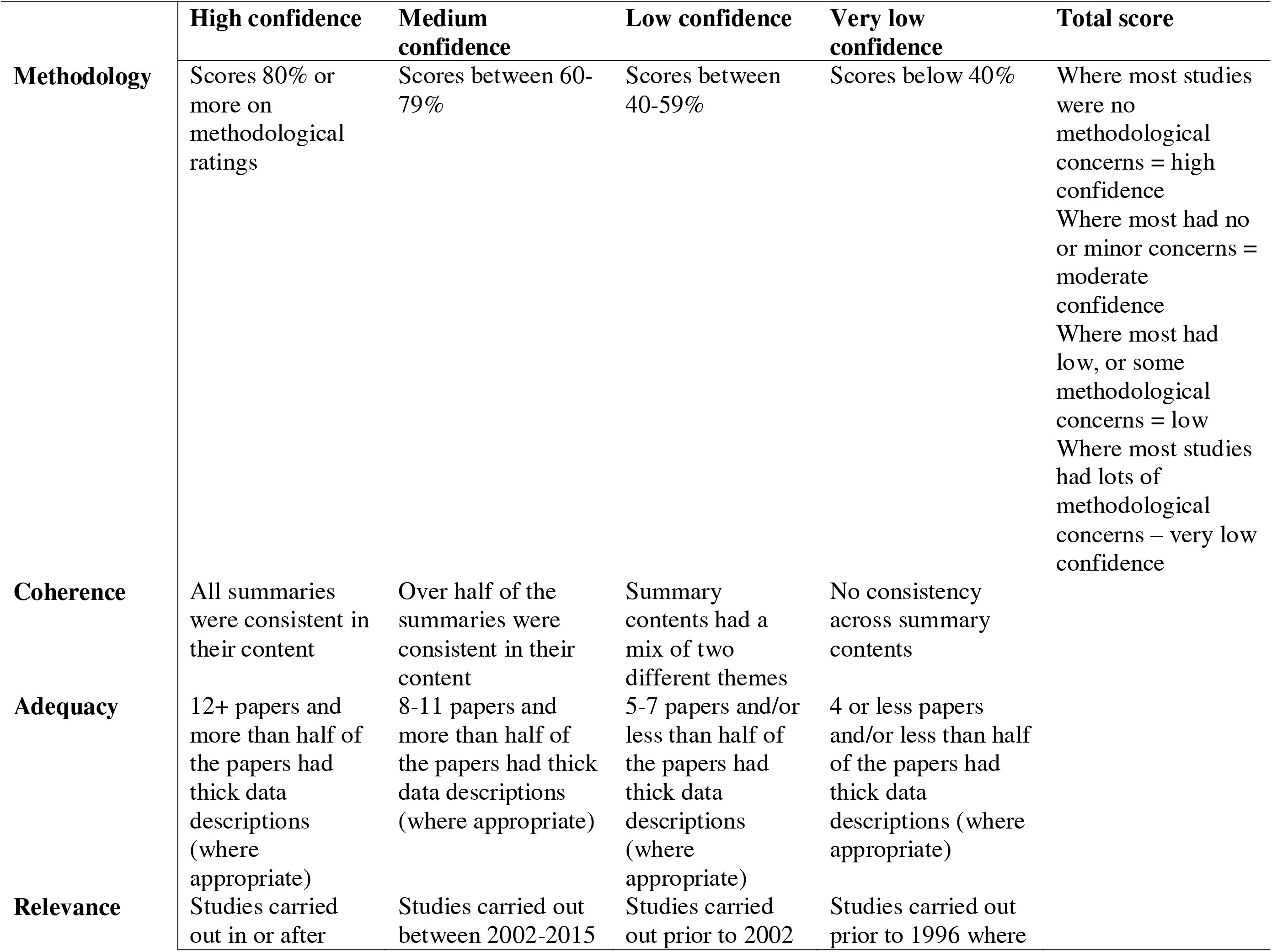
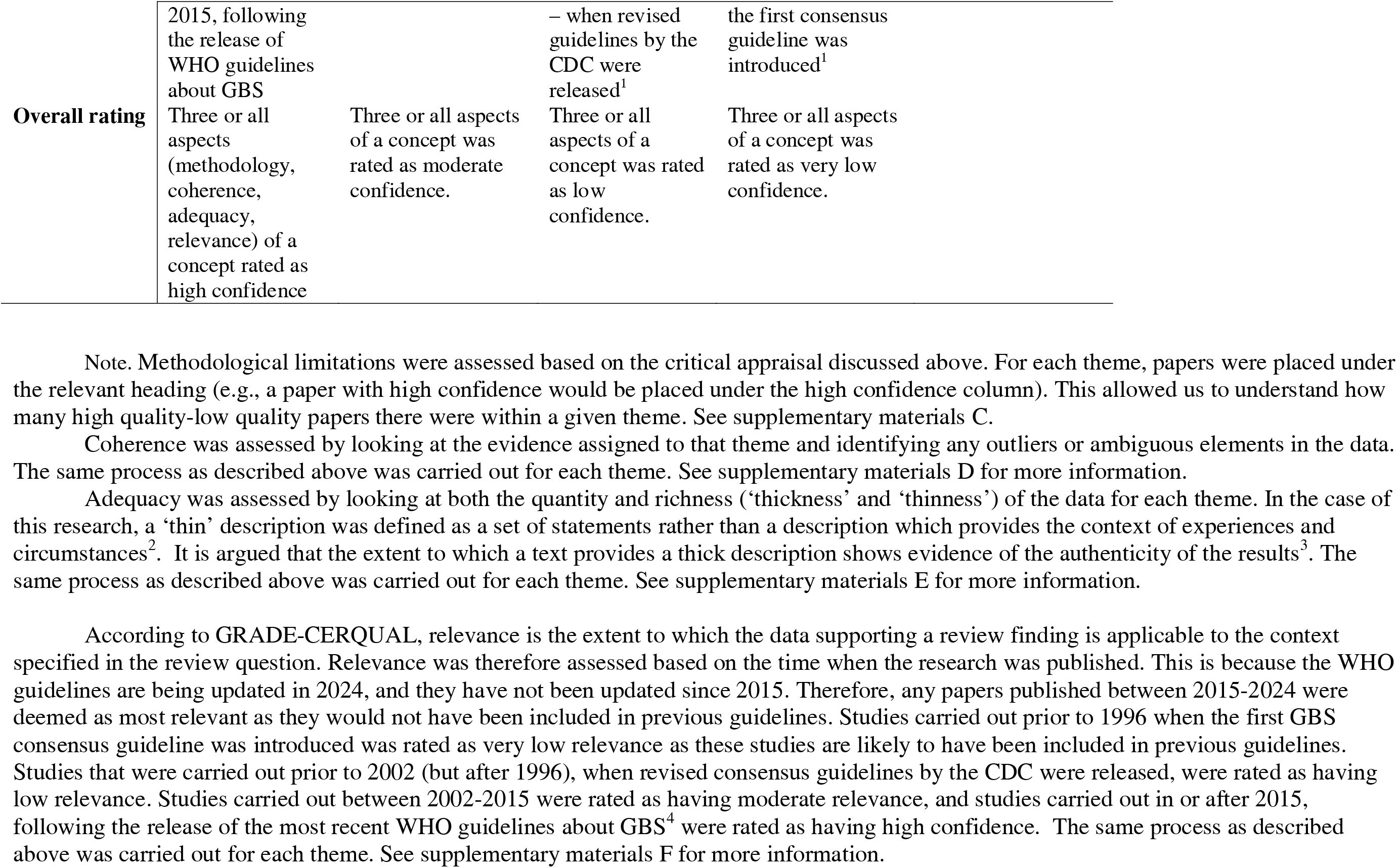
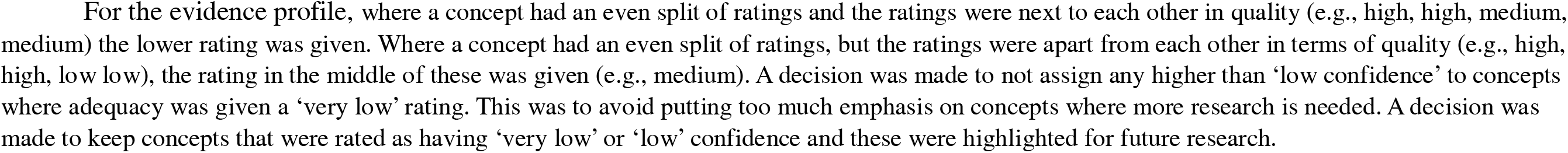
GRADE-CERQUAL Rules

## Results

### Study selection

Searches identified 2,096 papers and an additional 6 were identified by forward and backward searching. After 818 duplicates were removed, title and abstract screening excluded a further 1,205 papers. 74 full-text papers were screened by full text, as five were conference abstracts so did not have full texts. Of the 74 papers screened by full-text, 30 were excluded (see Figure 1).

**Figure 1.**
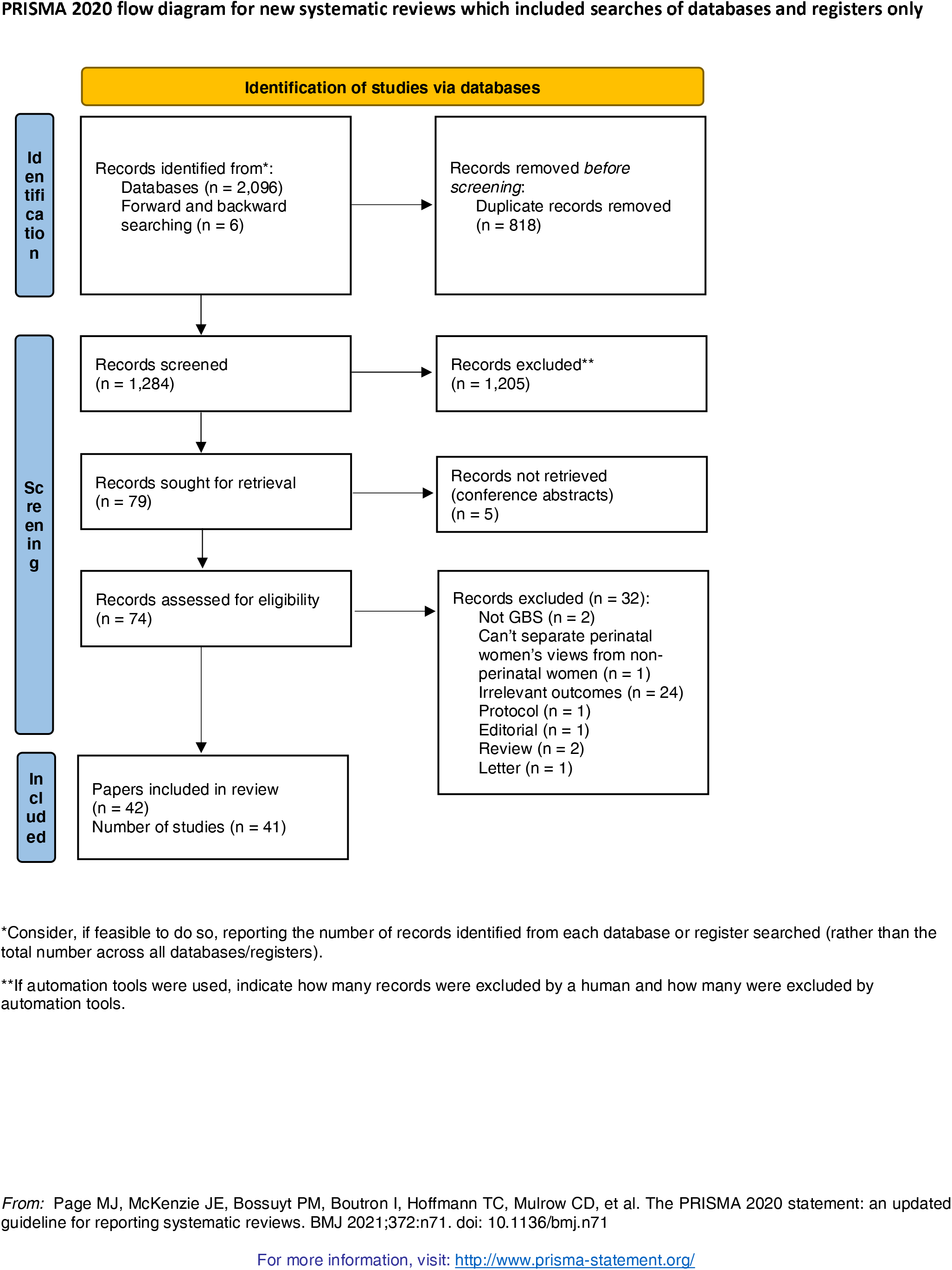
Prisma Flow Diagram

The reasons for exclusion included: the papers were not about GBS (n = 2); perinatal and non-perinatal women’s views could not be separated (n = 1); outcomes not relevant to the review (n = 22); protocol, editorial, review or letter (n = 5).

### Study characteristics

The review included 42 papers reporting findings from 41 studies (see Table 2). The majority of papers were quantitative (n = 33), followed by qualitative (n = 5) and mixed methods (n = 4).

**Table 2.**
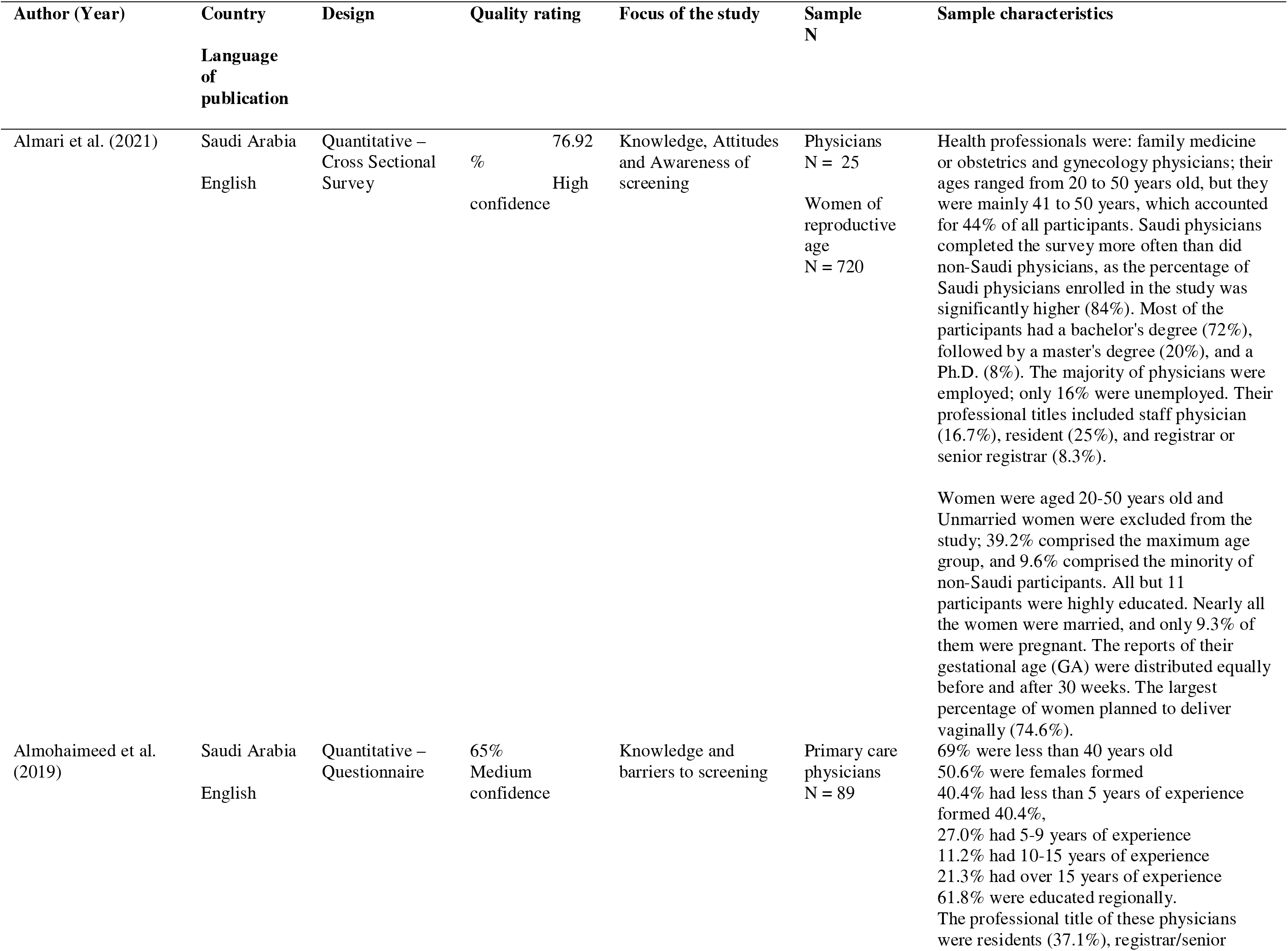
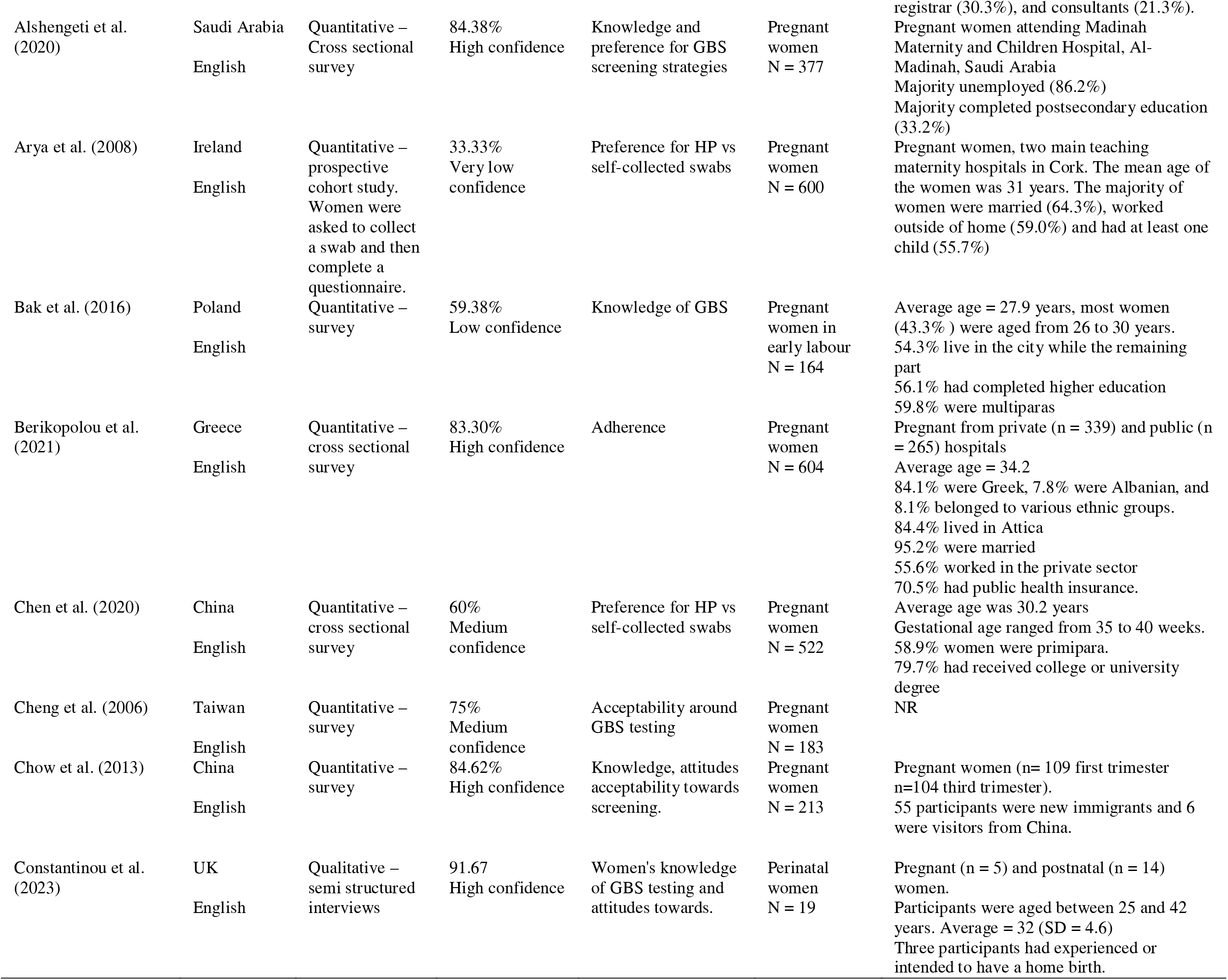
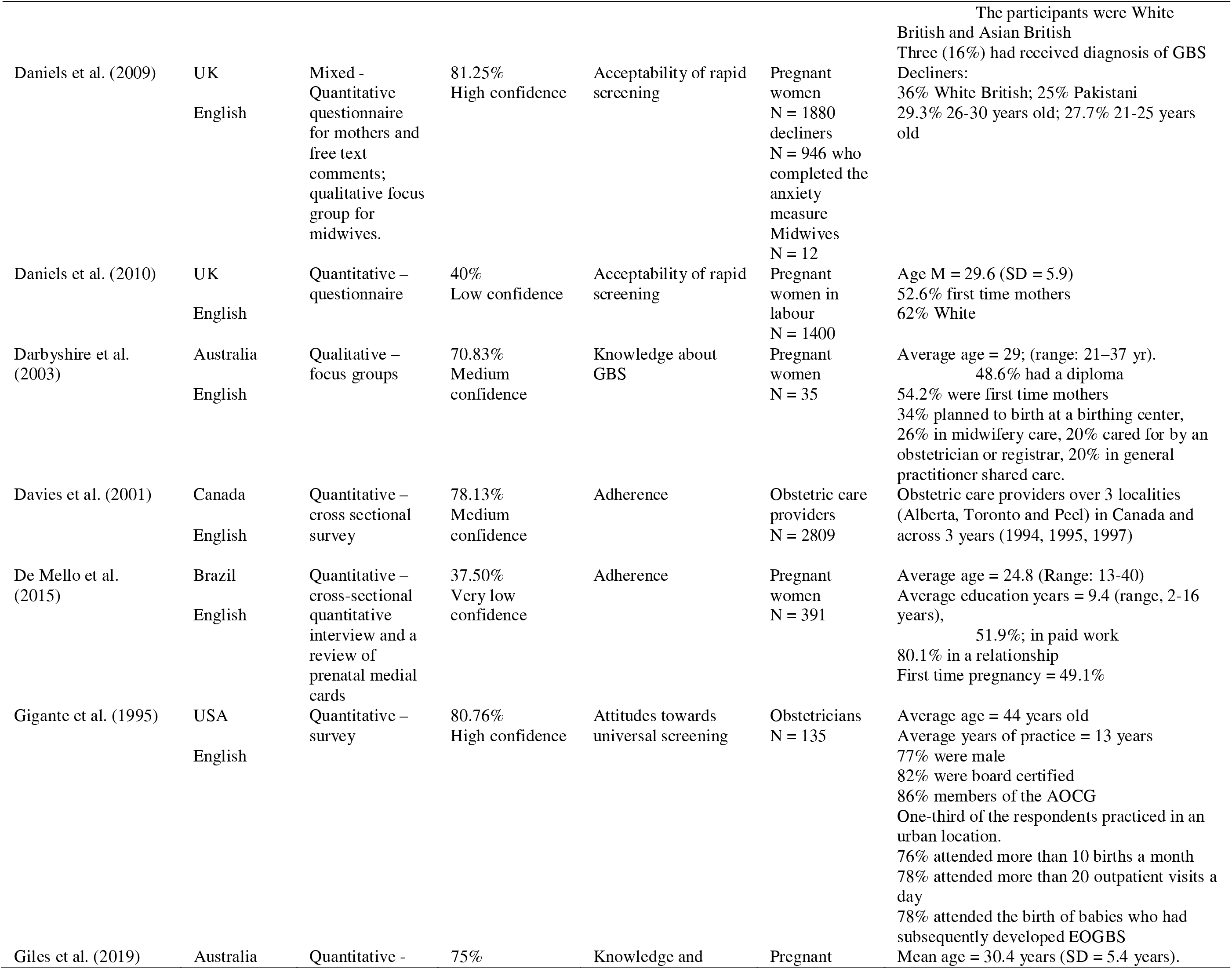
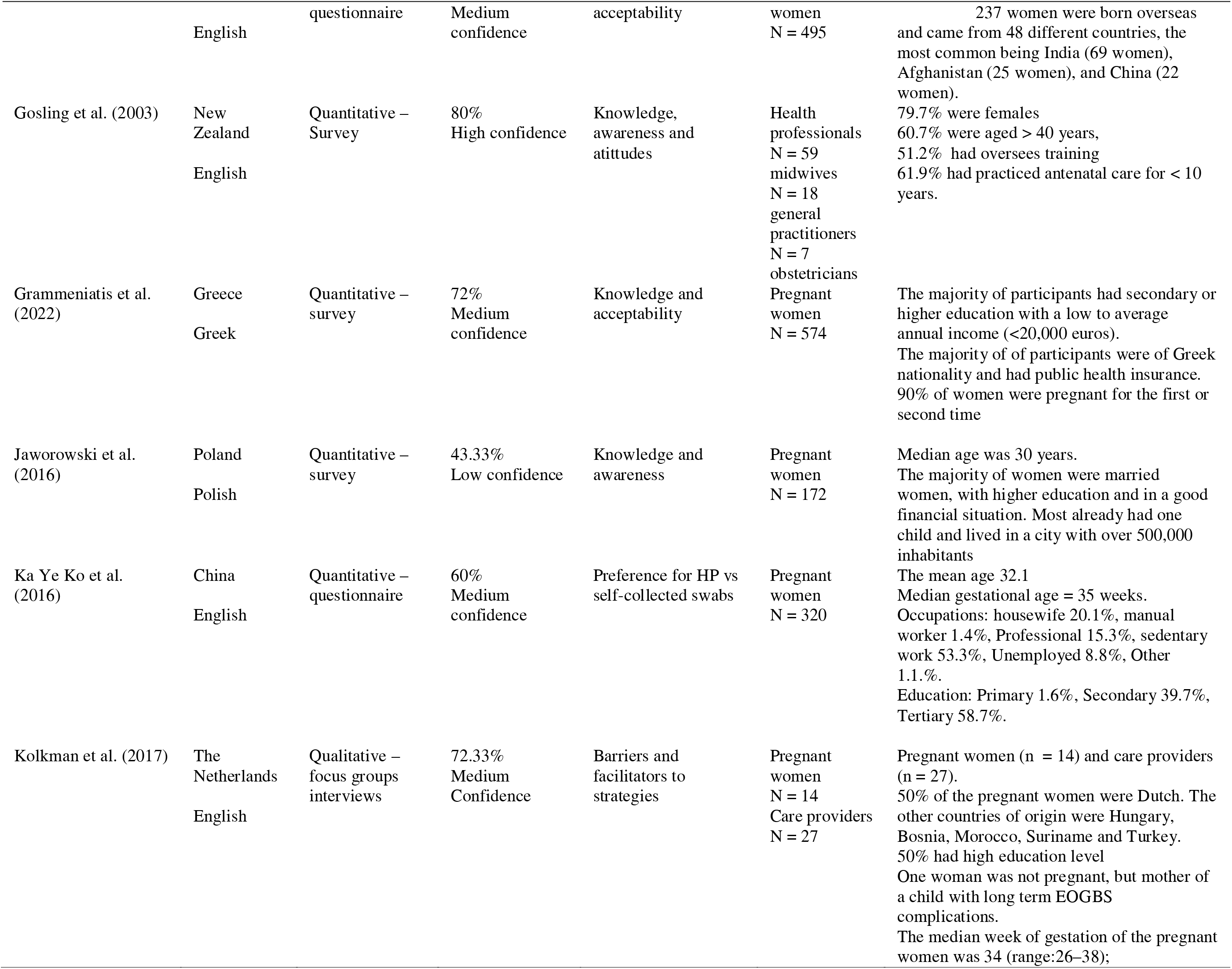
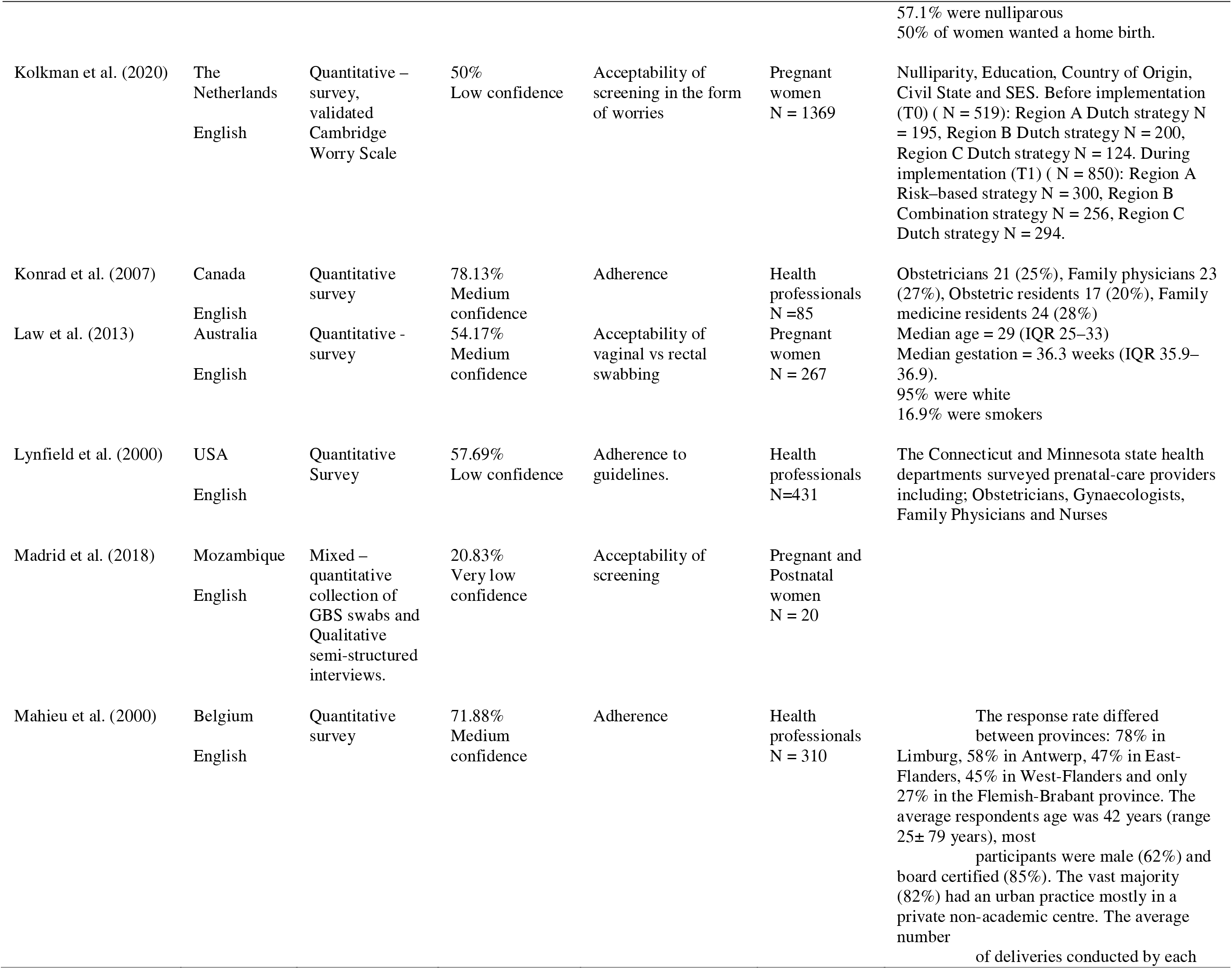
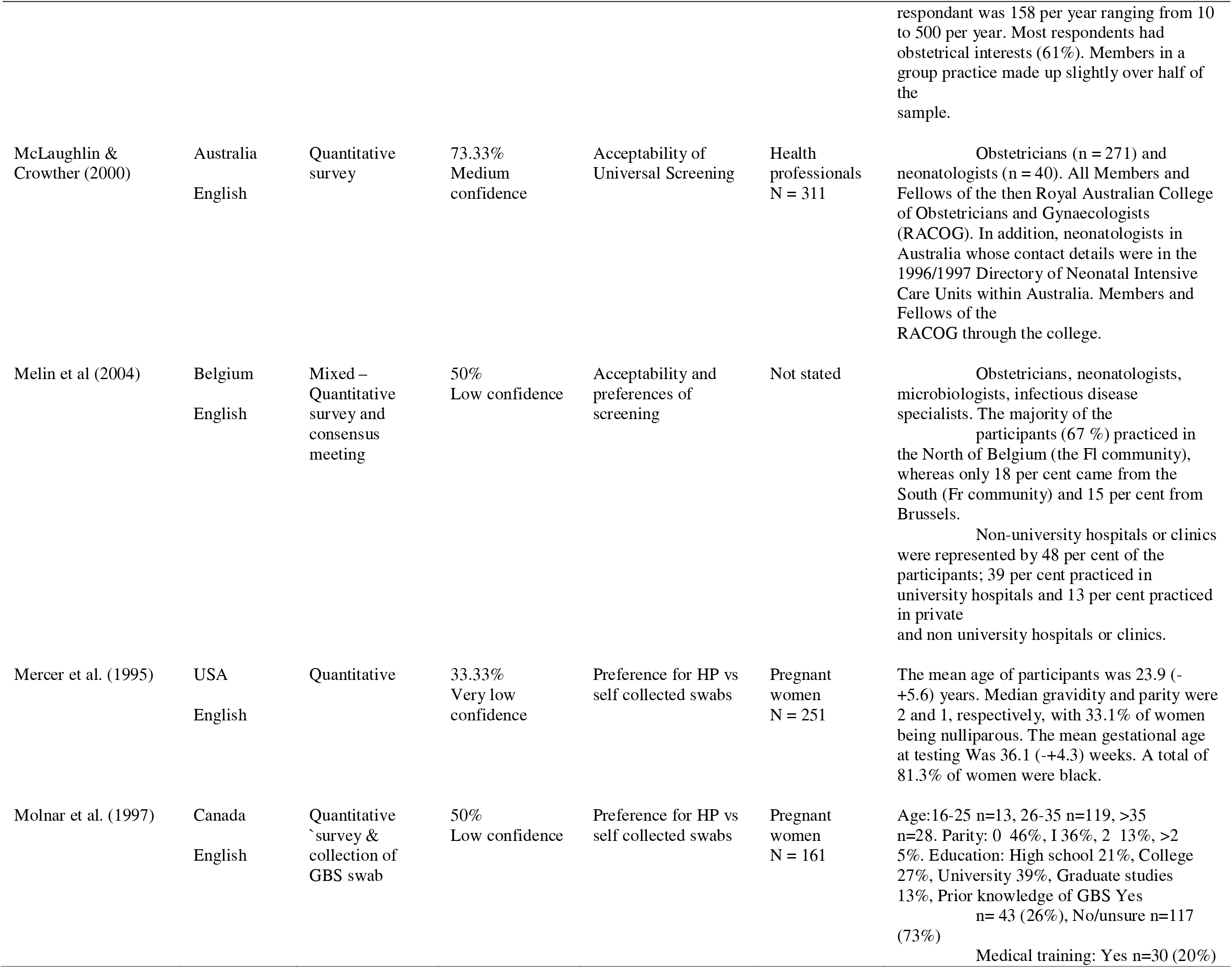
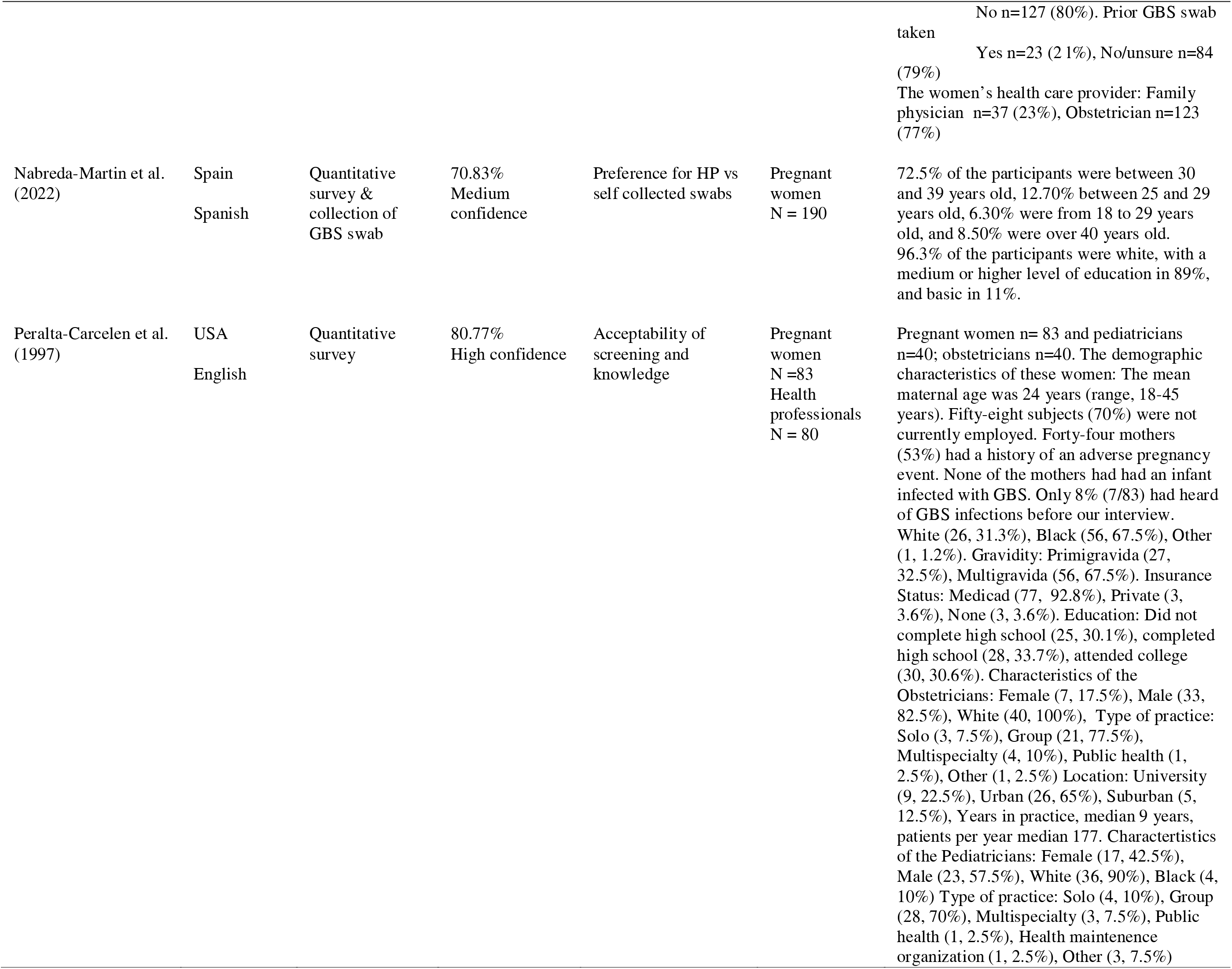
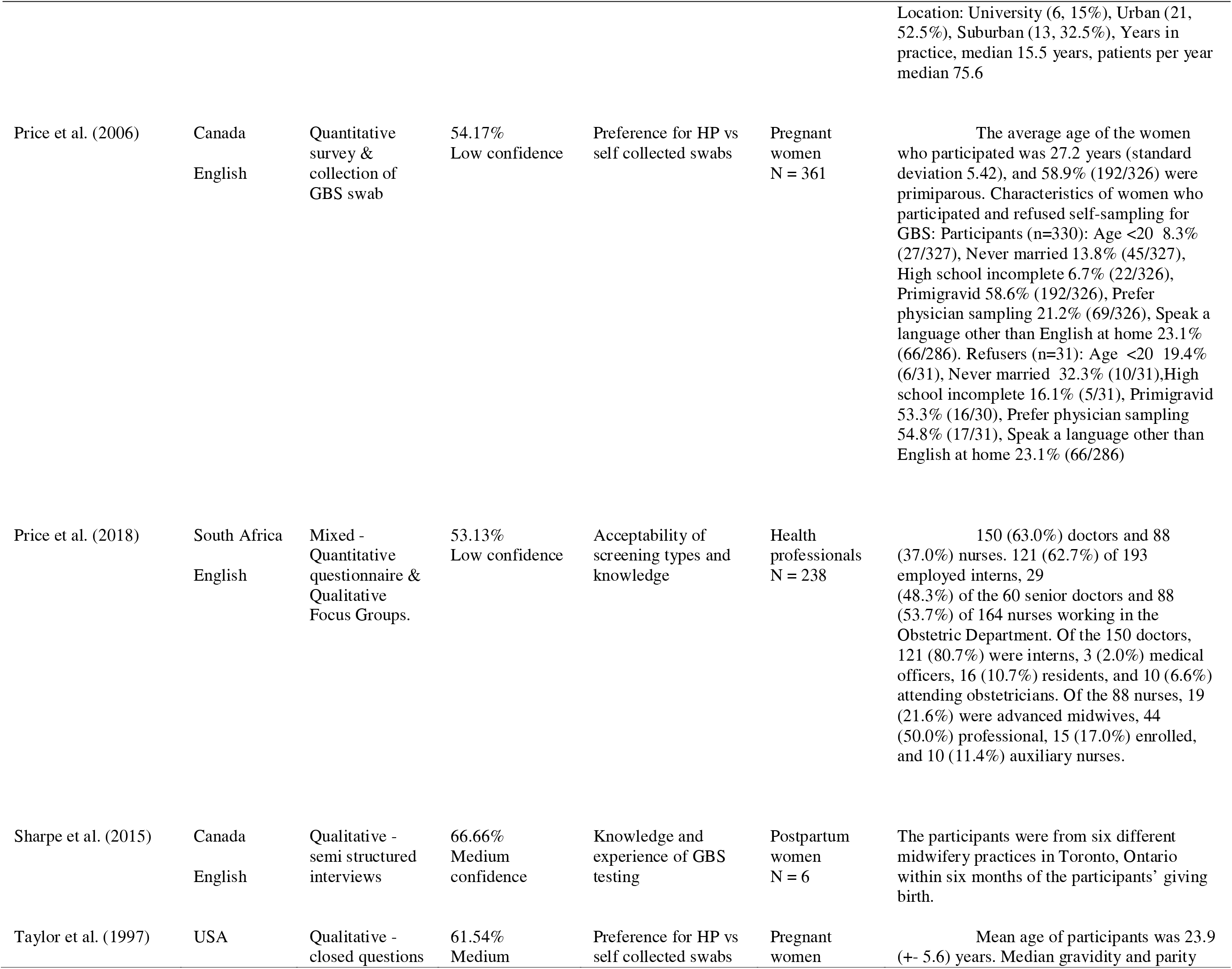
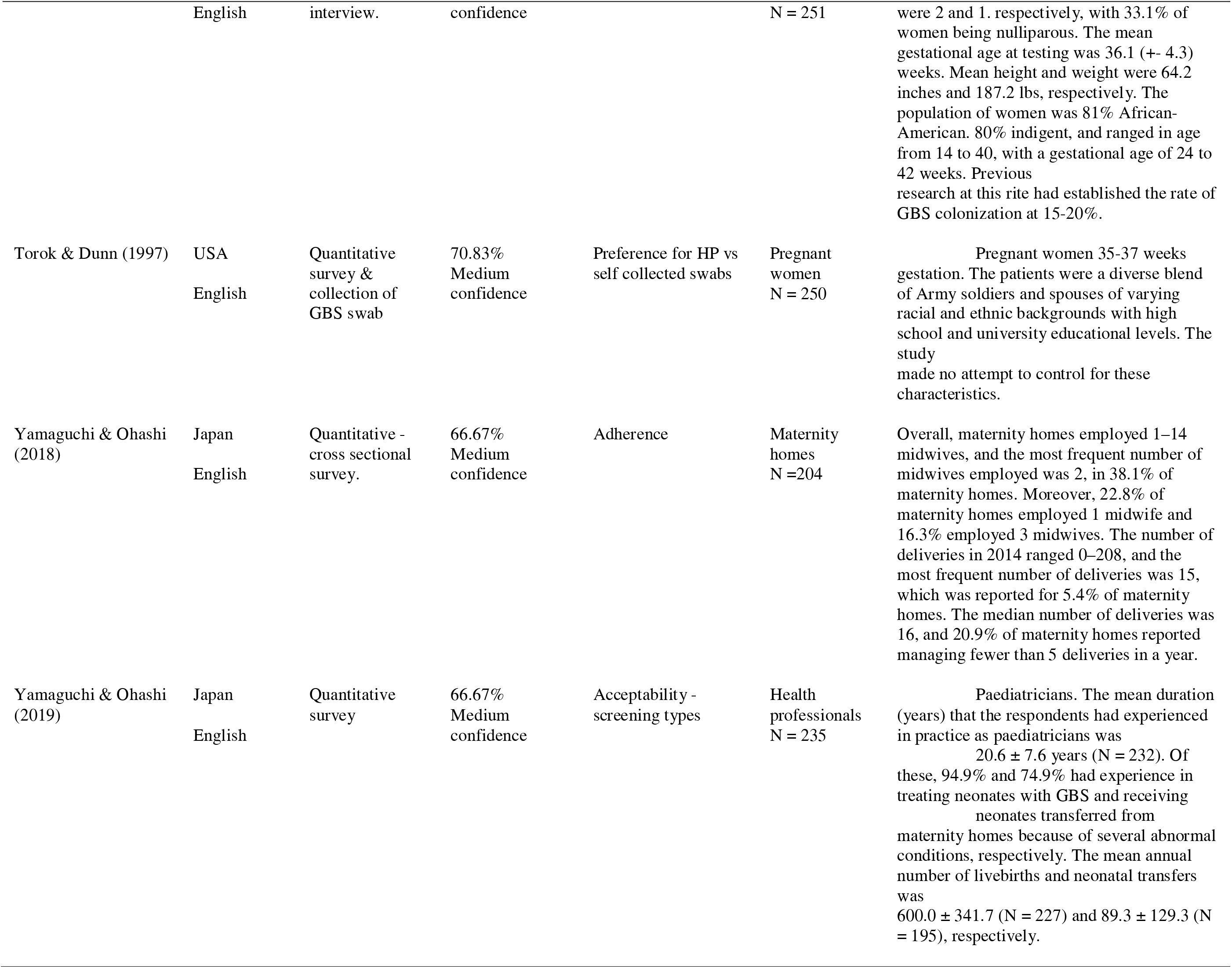
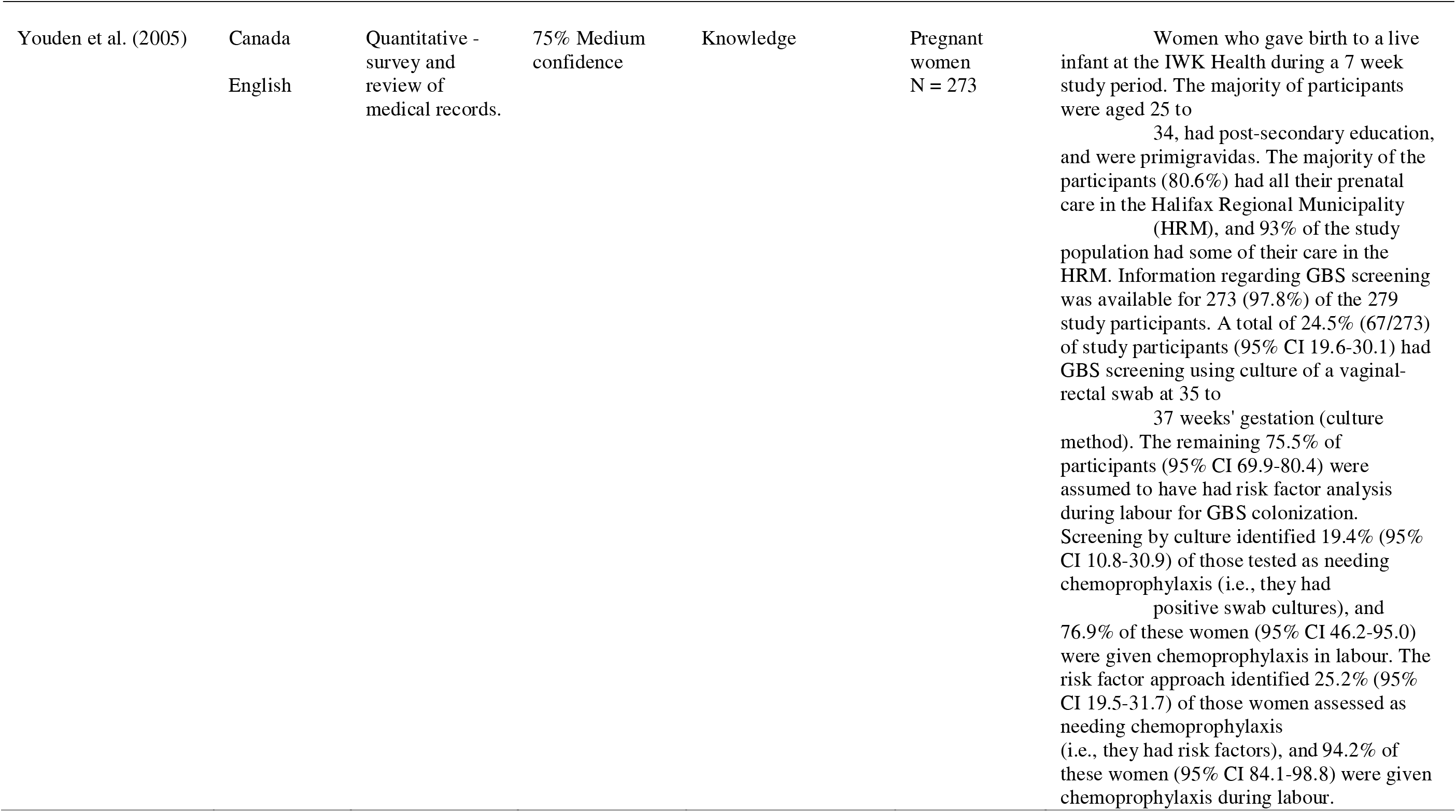
Study characteristics

Studies were conducted between 1995 – 2023 (Mean (M) = 2010; Median (Mdn) = 2013; Inter Quartile Range (IQR) = 2002-2018). The sample size varied from 6 to 2,809, with a total of 16,306 participants (M = 398; Mdn = 251; IQR = 163-431). Papers recruited health professionals (n = 12), pregnant and postnatal women (n = 26) or both women and health professionals (n = 4). The studies were conducted in 18 countries, with most being high income countries (Australia: n = 4; Canada: n = 6; USA: n = 6). Only one study was carried out in a lower-middle income country^31^ (Mozambique) and five were carried out in upper-middle income countries (Brazil: n = 1; China: n = 3; South Africa: n = 1). Three papers were not published in English (Greek, Polish, and Spanish) and were translated using Google translate.

### Risk of bias within studies

Fifty percent (n = 21) were rated as having medium confidence with the methodology; 8 were rated as high confidence; 9 were low confidence and 4 were rated as very low confidence with the methodology.

### Synthesis of results

Results were grouped according to the aims of the review, which led to 4 categories (Knowledge and awareness, Preferences, Acceptability, Feasibility/Adherence). Thirty-nine statements of findings were generated (see Table 3). Only statements of findings with high and moderate confidence will be discussed here. The remaining statements of findings can be found in Table 3.

**Table 3.**
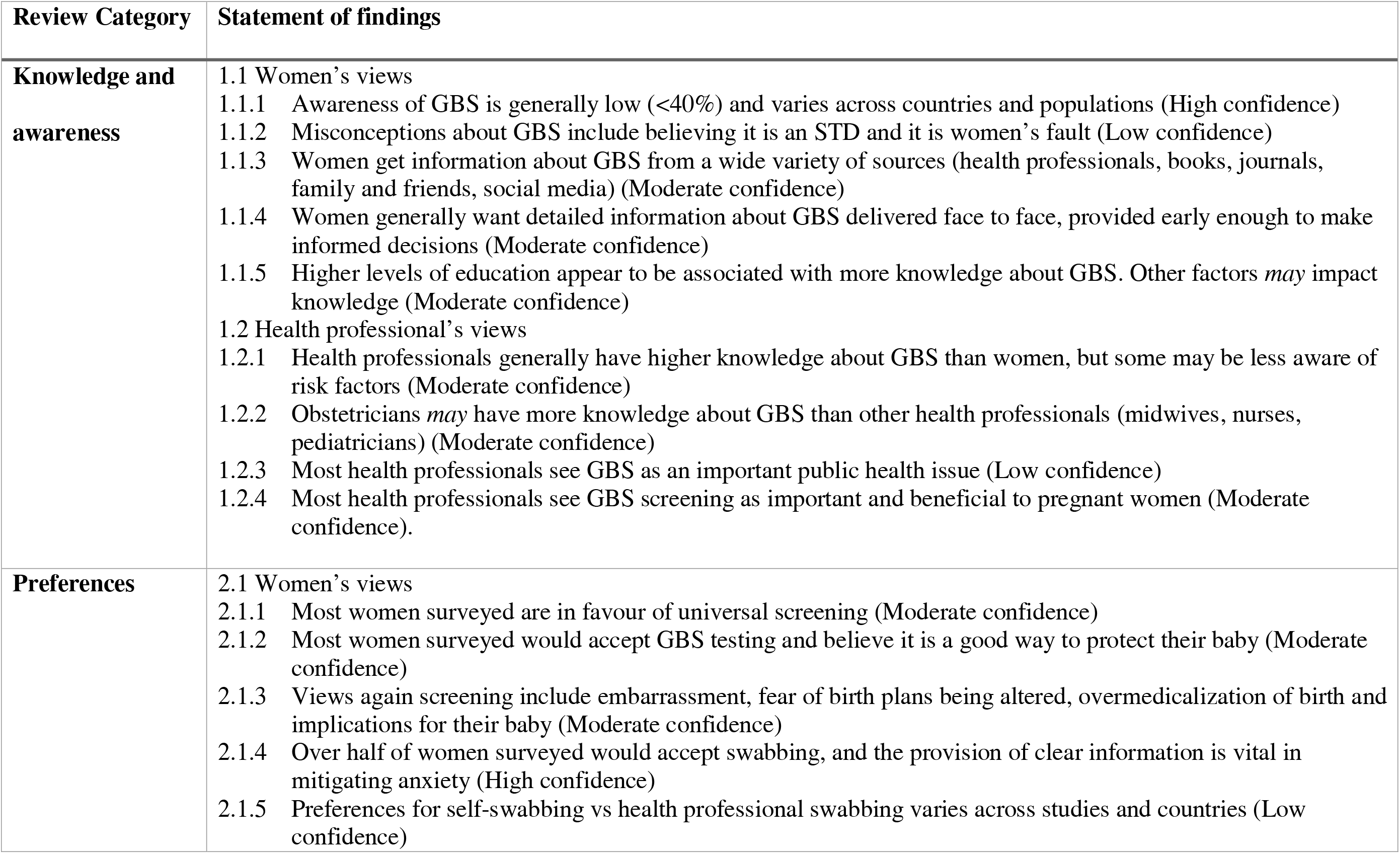
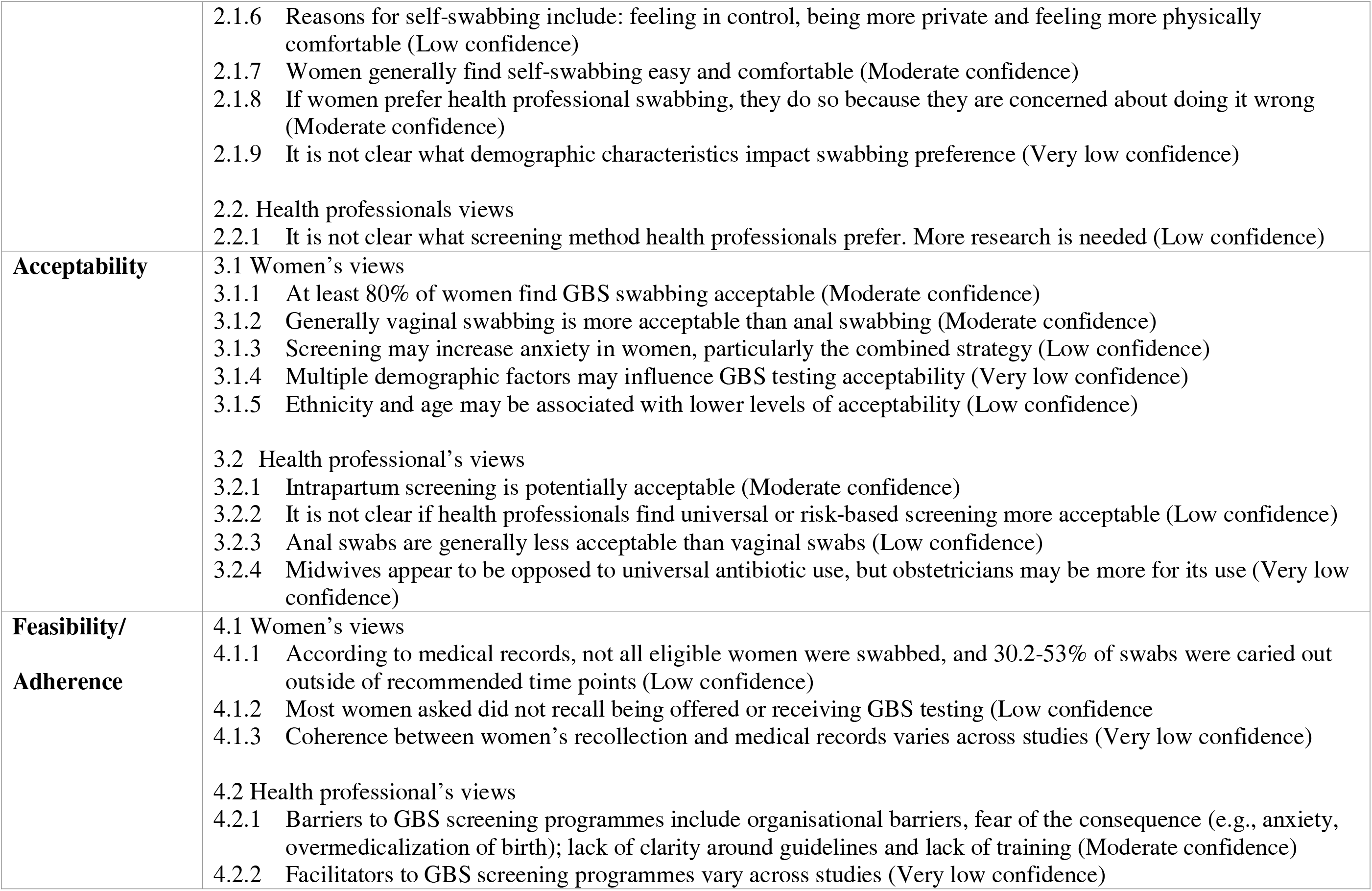
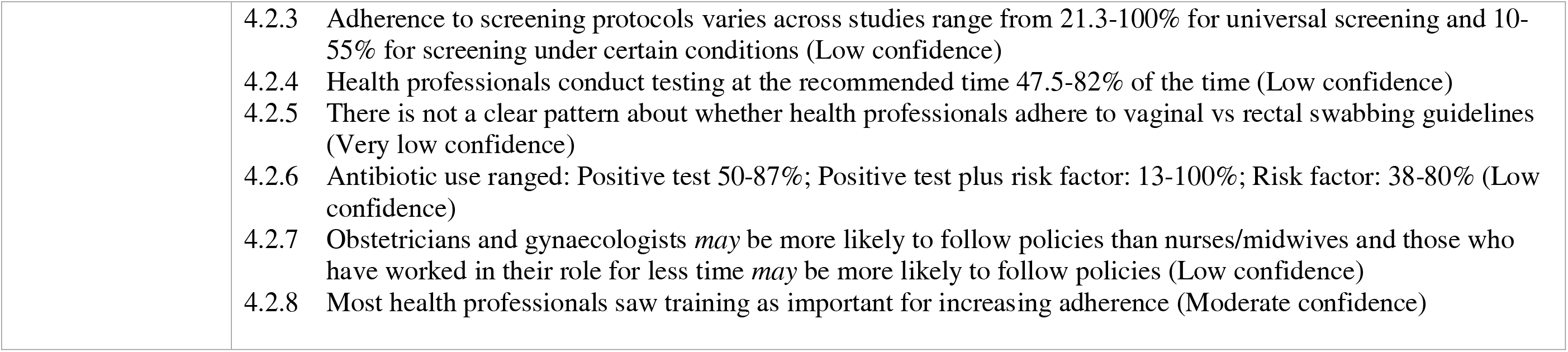
Categories and statement of findings

## Knowledge/Awareness

### Women’s views

#### Awareness of GBS is generally low (<40%) and varies across countries and populations (High confidence)

In included studies, awareness of GBS ranged from 8-37% (n = 6; M = 24.78; SD = 9.77)^32–37^. Awareness of GBS screening programmes ranged from 9 – 67.1% (n = 3; M = 44.27; SD = 25.30)^32, 38, 39^. This variable knowledge about GBS was corroborated by the qualitative studies which reported most women did not have an extensive knowledge about GBS^40–42^.

#### Women get information about GBS from a wide variety of sources (moderate confidence)

These sources include: health professionals^32, 39, 42^, social media/online^32, 40^, books or booklets^32, 41^, their work^40^, family and friends^40, 41^, personal experience^40, 41^, and antenatal education^40^.

#### Women generally want detailed information about GBS delivered face-to-face (Moderate confidence)

Women reported that they would like a range of information about GBS/GBS screening programmes in the form of leaflets, websites, and face-to-face detailed explanations. Positive GBS screening results should be delivered face-to-face^8, 40^. Some women felt the information they were given was inconsistent, unclear, poorly explained or inconsistently delivered.^32, 39, 42^

Higher levels of education appears to be associated with more knowledge about GBS. Other factors *may* impact knowledge (moderate confidence).

Five of seven quantitative studies found that the higher women’s education level, the more knowledge they had about GBS/GBS screening programmes^34–36, 38, 43^. Three studies looked at the impact of wealth and found that those with higher income tended to be more knowledgeable ^34, 36, 44^. Data suggest that more time spent in the country women are residing in can impact knowledge^,34, 35^, however this was only supported by two studies. No consistent relationship between knowledge and age^32, 35, 38, 43, 45^; employment status^32, 34, 35^; city vs village living^38, 44^; past exposure to GBS^32, 40, 44, 45^; or parity^32, 35, 38, 44, 45^ was found.

### Health professional’s views

#### Health professionals generally have higher knowledge about GBS than women (moderate confidence)

GBS screening knowledge is higher in health professionals with over 75% of health professionals sampled having good or excellent knowledge about GBS screening^32, 46^. Two studies looked at management strategies for identifying and treating GBS and found that at least 80% could identify a screening strategy.^47, 48^

#### Obstetricians *may* have more knowledge than other health professionals (moderate confidence)

Three quantitative studies report obstetricians tend to have more knowledge about GBS screening, management strategies and risk factors than midwives and paediatricians.^37, 48, 49^ One study found that older obstetricians had more knowledge than younger ones, but more research is needed^46^.

#### Most health professionals see screening as important and beneficial to women (moderate confidence)

Three studies found that most health professionals (between 69-91.1% of those sampled) thought screening for GBS was important^32, 46, 49^ and two studies found most health professionals sampled (72-88.8%) believed screening to be beneficial for pregnant women.^32, 46^

### Preferences

The review identified information about preferences in terms of women and health professional’s preference towards a specific GBS screening strategy, as well as women’s views in favour of screening, views against screening and preferences regarding swabbing itself.

### Women’s views

#### Most women surveyed are in favour of universal screening (moderate confidence)

Three studies ^34, 37, 45^ looked at universal based screening, and the majority of women (61.8%-81%) preferred this strategy. On the other hand, Kolkman et al. (2017) found that 86% of women preferred the combination strategy.

#### Most women surveyed would accept GBS testing and believe it is a good way to protect their baby (moderate confidence)

Two studies (one quantitative^34^ and one qualitative^40^) found that most women asked would be happy to accept GBS testing. Most women believed that testing was beneficial because it was a good way to protect their baby.^40^ The fact EOGBS can be prevented through antibiotics was seen as a positive thing, but clear information about it should be provided.^8, 40, 42^

#### Views against screening include embarrassment, fear of birth plans being altered, overmedicalization of birth and implications for their baby (moderate confidence)

Four studies reported negative views around testing. A small proportion of women in De Mello et al.’s sample reported feeling embarrassed or afraid after testing.^39^ Women were concerned about accepting screening because of the risk of increased stress and anxiety,^40^ risk of over treatment^8^, risk of over medicalising birth and ruining birth plans^40, 42^, potential negative effects for the woman and her baby are the safety of swabbing and antibiotic prophylaxis.^40, 42^

#### Over half of women surveyed would accept swabbing, and the provision of clear information is vital in mitigating anxiety (high confidence)

Two studies found the majority of women would accept vaginal swabbing^8, 34^. However, Chow et al. (2013)^34^ found that women were less likely to accept high vaginal swabbing (only 30% would accept) and anal swabbing (only 13% would accept). Qualitative data found that most women saw the swabs as not particularly intrusive^40, 42^, but would want to be provided with high quality information about what it involved so they could make an informed choice ^40^. The qualitative data also indicated that a lot of the anxiety surrounding testing and positive test results could be mitigated through clear explanations and information provided by health professionals^40, 42^.

#### Women generally find self-swabbing easy (Moderate confidence)

Four studies explored ease of self-swabbing, and all found that the majority of women found it to be easy.^50–53^

#### Preference for health professional swabbing is because some women are concerned about doing it wrong (Moderate confidence)

Eight studies reported reasons women gave for preferring health professional swabbing. Reasons given were: fears of doing it incorrectly^33, 42, 51, 53–55^; women’s concerns about the accuracy of the swab;^8, 50^ the belief that health professionals have more knowledge of swabbing;^50, 51^ physical difficulties performing the swab e.g., bump getting in the way; ^50, 51, 53^ women generally not liking the idea of self-swabbing, or touching their genitals to perform the swab; ^50, 53, 54^ and concern about hurting their baby if they self-swabbed.^53^

## Acceptability

15 studies looked at acceptability of GBS screening programmes to women and health professionals, anxiety around screening as well as facilitators and barriers to acceptability.

### Women’s views

#### At least 80% of women find GBS testing acceptable (moderate confidence)

Five studies ^34, 50, 56–58^ looked at the levels of acceptability for women in being screened for GBS, and how the screening was performed, with acceptability ranging from 81-100% (M = 94; SD = 6.95).

#### Generally vaginal swabbing is more acceptable than anal swabbing (moderate confidence)

Three studies reported percentages related to the acceptability of vaginal vs rectal swabbing^34, 57, 59^. Acceptability for vaginal swabbing ranged from 62-90% (M = 78.13; SD = 11.82). Acceptability for rectal swabbing ranged from 13-84% (M = 55.7; SD = 30.72).

### Health Professionals views

**Intrapartum screening is potentially acceptable** (moderate confidence). Three studies looked at the acceptability of antenatal vs intrapartum screening and although most health professionals found antenatal screening more acceptable^47, 60^, the proportion of the health professionals saying they would use rapid testing if it became available, was clinically proven and effective varied across studies (5% and 47%). Furthermore, a qualitative study found that most midwives felt that rapid testing was acceptable and possible during labour. However, they prioritised safe labour and birth care, and stated that they would not take swabs if it was inappropriate. Practical issues were raised with rapid intrapartum testing in terms of the difficulty of multi-tasking on a busy labour ward, or the speed at which some women labour. These issues were linked to staff shortages and if rapid testing was to become part of routine practice midwives believed there should be a dedicated person to do it. On the other hand, some respondents said it may not be worth adding an additional task to be carried out during labour given the low levels of EOGBS^61^.

## Feasibility

Eight studies were identified that reported barriers and facilitators to screening programmes.

Ten studies were also identified that looked at adherence to GBS screening protocols and can therefore be used as a proxy for feasibility, as low adherence is likely to reflect low feasibility.

### Health Professional Views

#### Barriers to GBS screening programmes include organisational barriers, fear of the consequence (e.g., anxiety, overmedicalization of birth); lack of clarity around guidelines and lack of training (moderate confidence)

The most commonly cited barrier was related to the organisation in which they worked including supervisors not supporting the use of the GBS protocol, or time constraints^8, 32, 46, 48, 49, 62^. Another common barrier was fear of the consequences of screening and providing antibiotics^8, 32, 46, 49^ including concerns over maternal discomfort and anxiety^8^, the risk of over treatment, antibiotic resistance, over medicalisation of birth and a reduction of the choice for women to home birth^8, 49^. Other reasons given were lack of clarity around the guidelines^8, 62^, medico- legal/political reasons^8, 62^; and lack of training^32, 46^

#### Most health professionals saw training as important for increasing adherence (moderate confidence)

Training in GBS was seen as important to improve adherence to GBS screening programmes.^32, 46, 48^ In a qualitative study, health professionals stated that engagement with GBS protocols could be encouraged by receiving feedback regarding the wellbeing of a neonate that had been affected by GBS infection, as this could sensitise them to the issue. They also stated that campaigns and media information focused on GBS could be important in improving engagement.^48^

## Discussion

The aim of this rapid review was to collate and synthesise the relevant evidence regarding GBS screening and provide a critical appraisal and overview of the evidence-base. The review looked at knowledge and awareness, preferences, acceptability and feasibility/adherence of GBS screening programmes for women and health professionals and identified a total of 4 categories and 39 statements of findings.

## Main findings

One of the statement of findings that had high confidence in the evidence was women’s low knowledge about GBS screening programmes. This is likely due to many reasons, including a lack of public awareness of GBS. Hunt (2012) speculated that this poor knowledge may be influenced by screening programmes generally being risk-based, rather than universal, meaning many midwives are not fully informed about GBS themselves, making it difficult to advise women in their care.^63^ This suggestion is supported by this review which found that obstetricians tend to have higher knowledge of GBS and may be more likely to discuss, screen for, and follow policies related to GBS than nursing and midwifery professionals^32, 37, 48, 49, 64^.

The other statement of findings that had high confidence in the evidence was that women’s attitudes are mostly, but not universally, positive towards GBS testing procedures, and that women generally prefer the universal based screening strategy (moderate confidence)^34, 37, 45^. This finding is consistent with other research which has found that women tend to find GBS vaccinations acceptable.^65^ A previous systematic review found that women value maintaining a healthy pregnancy for themselves and their baby.^66^ This may make testing more acceptable, as it could mean women feel they are mitigating risk and ensuring a healthy baby.^40^

However, this review also highlighted the importance of considering issues with over medicalising labour and birth, and in some cases iatrogenic harm for some women and babies in the case of antibiotic prophylaxis. Some women wish to focus on more of a holistic model of pregnancy and birth, with a view to minimise medical procedures as much as possible.^67^ This is supported by the statement of findings that some women are against testing due to stress and anxiety it could cause to the mother and the baby, the risk of over-medicalising birth, preventing home birth and the safety of swabbing and antibiotic treatment for the mother and their baby.^8, 40, 42^

## Strengths and limitations

The strengths of this rapid review are that it synthesises a large amount of information from 42 papers and used the CERQual^27^ approach to grade confidence with the evidence. This information can therefore be used to identify recommendations for the design and delivery of care^69^. In addition, no papers were excluded based on the language they were published in, meaning papers from 18 different countries were included. A limitation is that only papers published in academic journals were included. Relevant papers from health services, charities, third sector organisations and other grey literature may have been missed. Another limitation is that a score-based approach was used to categorise studies risk of bias into low, medium and high confidence, rather than a domain based approach^20^. This was done to allow for comparisons across studies.

## Interpretation

Women’s low knowledge about GBS suggests women need to be provided with high quality information regarding GBS, GBS screening procedures and antibiotic prophylaxis during antenatal care, or through antenatal education. Providing women with information about this will enable them to make informed decisions about their care. Health professionals may require more training on GBS screening to ensure they can provide the high quality information to women that they need.^40^ This is supported by two of the studies from the review which found that health professionals would like more training^32, 46^, and that a lack of training was seen as a barrier to implementing GBS screening procedures.

Women’s generally positive attitudes towards the GBS testing procedure suggest that most women would be happy to be swabbed for GBS. However, the review also identified concerns around a positive GBS result impacting women’s birth plans, and some concerns about the potential over-use of antibiotics. Given the importance of women’s birth plans being met in terms of increased birth satisfaction and reduced birth trauma and post-traumatic stress symptoms^68^ it is important for health professionals and service managers to weigh up the positives of screening for GBS, whilst also taking into account women’s individual values and birth plans.

Overall, half of the statement of findings were rated as having low confidence or very low confidence with the evidence (n = 21). This suggests more high-quality research is needed that examines women and health professionals’ views on knowledge, preferences, acceptability, and feasibility of GBS screening. There was no research on equity of delivering GBS screening. More research should therefore be carried out to identify what women and health professionals’ values surrounding GBS screening and treatment are, and what the key equity issues might be. Furthermore, the studies were carried out in 18 countries reflecting a very broad range of medical practice, social values, beliefs and medico-legal environments. It is likely that the heterogeneity may have impacted the results, therefore future research should focus on cross-cultural comparisons of knowledge, preferences, acceptability and adherence for GBS screening programmes.

## Conclusion

The aim of this rapid review was to synthesise evidence on women and health professionals’: (1) knowledge and awareness of; (2) preferences for; and (3) acceptability of GBS screening programmes, and (4) how feasible they are to implement. This is the first review that has been carried out that looks at women and health professionals views related to GBS screening preferences. Overall, only two statements of findings were rated as having high quality with the evidence and these were women’s low knowledge of GBS and GBS screening programmes, and most women’s generally positive attitudes towards swabbing for GBS. The results from the review suggest that women should be provided with high quality information about GBS and GBS screening programmes in order to make informed decisions about their care. Health professional training may need to be increased in order to do this. Furthermore, it is important for health professionals and service managers to weigh up the positives of screening for GBS, whilst also taking into account women’s individual values and birth plans. More research is needed on this topic, specifically around the equity and feasibility of implementing GBS screening programmes.

## Data Availability

All data produced in the present study are available upon reasonable request to the authors

