## Supplemental Material A for "Acceptability and feasibility of maternal screening for Group B Streptococcus: a rapid review"

Supplementary material

A- Search Syntax

| **Syntax** | **Database** | **Hits** |
| --- | --- | --- |
| (Women OR Mother OR Parent OR Father OR Partner OR "Health care professional*" OR "Health professional*" Obstetrician* OR Midwi* OR Maternity) AND (Pregnan* OR *natal OR *partum OR Birth* OR Delivery OR Childbirth OR Labour/Labor) AND (GBS OR "Group B Strep" OR "Group B Streptococcus" OR "Group B Streptococcal" OR "GBS Bacteria" OR "neonatal Infection" OR "neonatal Sepsis" OR "neonatal Pneumonia" OR "neonatal meningitis") AND (Test* OR Screen* OR Sampl* OR Swab* OR "Routine testing" OR "Universal testing" OR "Risk-factor approach" OR Vaccin*) AND (Value OR View OR Experience OR Preference OR Perspective OR Knowledge OR Attitude OR Acceptab* OR Feasib* OR Equity OR Adherence OR Compliance) Limiters- Human | Medline | 587 |
| (Women OR Mother OR Parent OR Father OR Partner OR "Health care professional*" OR "Health professional*" Obstetrician* OR Midwi* OR Maternity) AND (Pregnan* OR *natal OR *partum OR Birth* OR Delivery OR Childbirth OR Labour/Labor) AND (GBS OR "Group B Strep" OR "Group B Streptococcus" OR "Group B Streptococcal" OR "GBS Bacteria" OR "neonatal Infection" OR "neonatal Sepsis" OR "neonatal Pneumonia" OR "neonatal meningitis") AND (Test* OR Screen* OR Sampl* OR Swab* OR "Routine testing" OR "Universal testing" OR "Risk-factor approach" OR Vaccin*) AND (Value OR View OR Experience OR Preference OR Perspective OR Knowledge OR Attitude OR Acceptab* OR Feasib* OR Equity OR Adherence OR Compliance) Limiters - Population Group: Human | PsycINFO | 11 |
| (Women OR Mother OR Parent OR Father OR Partner OR "Health care professional*" OR "Health professional*" Obstetrician* OR Midwi* OR Maternity) AND (Pregnan* OR *natal OR *partum OR Birth* OR Delivery OR Childbirth OR Labour/Labor) AND (GBS OR "Group B Strep" OR "Group B Streptococcus" OR "Group B Streptococcal" OR "GBS Bacteria" OR "neonatal Infection" OR "neonatal Sepsis" OR "neonatal Pneumonia" OR "neonatal meningitis") AND (Test* OR Screen* OR Sampl* OR Swab* OR "Routine testing" OR "Universal testing" OR "Risk-factor approach" OR Vaccin*) AND (Value OR View OR Experience OR Preference OR Perspective OR Knowledge OR Attitude OR Acceptab* OR Feasib* OR Equity OR Adherence OR Compliance) Limiters- Human | CINAHL | 133 |
| (Women OR Mother OR Parent OR Father OR Partner OR "Health care professional*" OR "Health professional*" Obstetrician* OR Midwi* OR Maternity) AND (Pregnan* OR *natal OR *partum OR Birth* OR Delivery OR Childbirth OR Labour/Labor) AND (GBS OR "Group B Strep" OR "Group B Streptococcus" OR "Group B Streptococcal" OR "GBS Bacteria" OR "neonatal Infection" OR "neonatal Sepsis" OR "neonatal Pneumonia" OR "neonatal meningitis") AND (Test* OR Screen* OR Sampl* OR Swab* OR "Routine testing" OR "Universal testing" OR "Risk-factor approach" OR Vaccin*) AND (Value OR View OR Experience OR Preference OR Perspective OR Knowledge OR Attitude OR Acceptab* OR Feasib* OR Equity OR Adherence OR Compliance) Limiters - Population Group: Human | PsychArticles | 45 |
| TITLE-ABS-KEY ( women OR mother OR parent OR father OR partner OR "Health care professional*" OR "Health professional*" obstetrician* OR midwi* OR maternity ) AND ( pregnan* OR *natal OR *partum OR birth* OR delivery OR childbirth OR labour/labor ) AND ( gbs OR "Group B Strep" OR "Group B Streptococcus" OR "Group B Streptococcal" OR "GBS Bacteria" OR "neonatal Infection" OR "neonatal Sepsis" OR "neonatal Pneumonia" OR "neonatal meningitis" ) AND ( test* OR screen* OR sampl* OR swab* OR "Routine testing" OR "Universal testing" OR "Risk-factor approach" OR vaccin* ) AND ( value OR view OR experience OR preference OR perspective OR knowledge OR attitude OR acceptab* OR feasib* OR equity OR adherence OR compliance ) AND ( LIMIT-TO ( EXACTKEYWORD , "Human" ) ) | SCOPUS | 236 |
| (Women OR Mother OR Parent OR Father OR Partner OR "Health care professional*" OR "Health professional*" Obstetrician* OR Midwi* OR Maternity) AND (Pregnan* OR *natal OR *partum OR Birth* OR Delivery OR Childbirth OR Labour/Labor) AND (GBS OR "Group B Strep" OR "Group B Streptococcus" OR "Group B Streptococcal" OR "GBS Bacteria" OR "neonatal Infection" OR "neonatal Sepsis" OR "neonatal Pneumonia" OR "neonatal meningitis") AND (Test* OR Screen* OR Sampl* OR Swab* OR "Routine testing" OR "Universal testing" OR "Risk-factor approach" OR Vaccin*) AND (Value OR View OR Experience OR Preference OR Perspective OR Knowledge OR Attitude OR Acceptab* OR Feasib* OR Equity OR Adherence OR Compliance) Filters: Humans | PubMed | 481 |
| tw:((women OR mother OR parent OR father OR partner OR "Health care professional*" OR "Health professional*" obstetrician* OR midwi* OR maternity) AND (pregnan* OR *natal OR *partum OR birth* OR delivery OR childbirth OR labour/labor) AND (gbs OR "Group B Strep" OR "Group B Streptococcus" OR "Group B Streptococcal" OR "GBS Bacteria" OR "neonatal Infection" OR "neonatal Sepsis" OR "neonatal Pneumonia" OR "neonatal meningitis") AND (test* OR screen* OR sampl* OR swab* OR "Routine testing" OR "Universal testing" OR "Risk-factor approach" OR vaccin*) AND (value OR view OR experience OR preference OR perspective OR knowledge OR attitude OR acceptab* OR feasib* OR equity OR adherence OR compliance)) AND ( mj:("Humans")) | Global Index Medicus | 8 |
| (Women OR Mother OR Parent OR Father OR Partner OR "Health care professional*" OR "Health professional*" Obstetrician* OR Midwi* OR Maternity) AND (Pregnan* OR *natal OR *partum OR Birth* OR Delivery OR Childbirth OR Labour/Labor) AND (GBS OR "Group B Strep" OR "Group B Streptococcus" OR "Group B Streptococcal" OR "GBS Bacteria" OR "neonatal Infection" OR "neonatal Sepsis" OR "neonatal Pneumonia" OR "neonatal meningitis") AND (Test* OR Screen* OR Sampl* OR Swab* OR "Routine testing" OR "Universal testing" OR "Risk-factor approach" OR Vaccin*) AND (Value OR View OR Experience OR Preference OR Perspective OR Knowledge OR Attitude OR Acceptab* OR Feasib* OR Equity OR Adherence OR Compliance) | Academic Search Ultimate | 324 |
| (Women or Mother or Parent or Father or Partner or "Health care professional*" or "Health professional* Obstetrician*" or Midwi* or Maternity).mp. and (Pregnan*.mp. or *natal/ or *partum/ or Birth*.mp. or Delivery.mp. or Childbirth.mp. or Labour 0R Labor.mp.) and (GBS or "Group B Strep" or "Group B Streptococcus" or "Group B Streptococcal" or "GBS Bacteria" or "neonatal Infection" or "neonatal Sepsis" or "neonatal Pneumonia" or "neonatal meningitis").mp. and (Test* or Screen* or Sampl* or Swab* or "Routine testing" or "Universal testing" or "Risk-factor approach" or Vaccin*).mp. and (Value or View or Experience or Preference or Perspective or Knowledge or Attitude or Acceptab* or Feasib* or Equity or Adherence or Compliance).mp. [mp=title, abstract, heading word, drug trade name, original title, device manufacturer, drug manufacturer, device trade name, keyword heading word, floating subheading word, candidate term word] limit 1 to (human and "remove medline records") | EMBASE | 261 |
| TI=((Women OR Mother OR Parent OR Father OR Partner OR "Health care professional*" OR "Health professional*" Obstetrician* OR Midwi* OR Maternity) AND (Pregnan* OR *natal OR *partum OR Birth* OR Delivery OR Childbirth OR Labour/Labor) AND (GBS OR "Group B Strep" OR "Group B Streptococcus" OR "Group B Streptococcal" OR "GBS Bacteria" OR "neonatal Infection" OR "neonatal Sepsis" OR "neonatal Pneumonia" OR "neonatal meningitis") AND (Test* OR Screen* OR Sampl* OR Swab* OR "Routine testing" OR "Universal testing" OR "Risk-factor approach" OR Vaccin*) AND (Value OR View OR Experience OR Preference OR Perspective OR Knowledge OR Attitude OR Acceptab* OR Feasib* OR Equity OR Adherence OR Compliance)) | Web of Science | 16 |
|  | **Total:** | 2,102 |
