## Supplemental Material C for "Acceptability and feasibility of maternal screening for Group B Streptococcus: a rapid review"

**CERQUAL Methodological quality scale**

| **Statement of findings** | **Definition** | **Studies citing this** | **N** | **No methodological concerns**  **(High confidence)** | **Minor concerns**  **(Moderate confidence)** | **Moderate concerns**  **(Low confidence)** | **Serious concerns**  **(Very low confidence)** | **Overall methodological confidence rating*** |
| --- | --- | --- | --- | --- | --- | --- | --- | --- |
| **Review Category: Knowlededge and awareness** | | | | | | | | |
| **1.1 Women’s views** | | | | | | | | |
| 1.1.1 Awareness   Awareness of GBS is generally low (<40%) and varies across countries and populations. | The percentage of women in the study sample with knowledge about GBS, GBS testing/screening and treatment for GBS. | Alamri et al., 2021; Alshengeti et al. 2020; Arya et al., 2008; Bak et al., 2016; Chow et al., 2013; Constantinou et al., 2023; Darbyshire et al., 2003; De Mello et al., 2015; Giles et al., 2019; Grammeniatis et al., 2022; Peralta-Carcelen et al., 1997; Sharpe et al., 2015; Youden et al., 2005 | 13 | Alshegenti et al. (2020)  Chow et al. (2013)  Constantinou et al. (2023)  Peralta-Carcelen et al. (1997) | Alamri et al. (2021)  Darbyshire et al. (2003)  Giles et al. (2019)  Grammeniatis et al. (2022)  Sharpe et al. (2015)  Youden et al. (2005) | Bak et al. (2016) | Arya et al. (2008)  De Mello et al. (2015) | **Moderate confidence –** most of the studies had no or minor methodological concerns |
| 1.1.2 Misconceptions about GBS   Misconceptions about GBS include believing it is an STD and it is women’s fault | Incorrect beliefs women may hold about GBS. | Alamri et al., 2021; Alshengeti et al. 2020; Chow et al., 2013; Constantinou et al., 2023; Darbyshire et al., 2003; Grammeniatis et al., 2022; Sharpe et al., 2015; | 7 | Alshegenti et al. (2020)  Chow et al. (2013)  Constantinou et al. (2023) | Alamri et al. (2021)  Darbyshire et al. (2003)  Grammeniatis et al. (2022)  Sharpe et al. (2015) |  |  | **Moderate confidence –** most of the studies had no or minor methodological concerns |
| 1.1.3 Where does women’s knowledge about GBS come from?   Women get information about GBS from a wide variety of sources (health professionals, books, journals, family and friends, social media) | Sources and locations that women access knowledge about GBS | Alamri et al., 2021; Constantinou et al., 2023; Darbyshire et al., 2003; De Mello et al. (2015); Sharpe et al., 2015; | 5 | Constantinou et al. (2023) | Alamri et al. (2021)  Darbyshire et al. (2003)  Sharpe et al. (2015) |  | De Mello et al. (2015) | **Moderate confidence –** most of the studies had no or minor methodological concerns |
| 1.1.4 What knowledge do women want?   Women generally want detailed information about GBS delivered face to face, provided early enough to make informed decisions. | Information that women would like to receive about GBS and how they would like to receive this information | Alamri et al., 2021; Constantinou et al., 2023; De Mello et al. 2015; Kolkman et al. 2017; Sharpe et al., 2015; | 5 | Constantinou et al. (2023) | Alamri et al. (2021)  Kolkman et al. (2017)  Sharpe et al. (2015) |  | De Mello et al. (2015) | **Moderate confidence –** most of the studies had no or minor methodological concerns |
| 1.1.5. Factors associated with knowledge  Higher levels of education appear to be associated with more knowledge about GBS. Other factors may impact knowledge | Demographic and health characteristics that might impact women’s GBS knowledge | Alamri et al., 2021; Alshengeti et al. 2020; Bak et al., 2016; Chow et al., 2013; Constantinou et al., 2023; Giles et al., 2019; Grammeniatis et al., 2022; Jaworowski et al., 2016; Youden et al., 2005 | 9 | Alshegenti et al. (2020)  Chow et al. (2013)  Constantinou et al. (2023) | Alamri et al. (2021)  Giles et al. (2019)  Grammeniatis et al. (2022)  Youden et al. (2005) | Bak et al. (2016)  Jaworowski et al. (2016) |  | **Moderate confidence –** most of the studies had no or minor methodological concerns |
| **1.2 Health professionals** | | | | | | | | |
| 1.2.1 Awareness  Health professionals generally have higher knowledge about GBS, but some may be less aware of risk factors. | The percentage of health professionals the study sample with knowledge about GBS, GBS testing/screening and treatment for GBS. | Alamri et al., 2021; Almohaimeed et al., 2019; Gosling et al., 2002; Melin et al. 2004; Peralta-Carcelen et al., 1997; Price et al., 2018; | 6 | Gosling et al. (2002)  Peralta-Carcelen et al. (1997) | Alamri et al. (2021)  Almohaimeed et al. (2019) | Melin et al. (2004  Price et al. (2018) |  | **Moderate confidence –** equal split across confidence levels, so have gone for the middle |
| 1.2.2 Factors associated with knowledge  Obstetricians *may*  have more knowledge about GBS than other health professionals (midwives, nurses, paediatricians). | Characteristics that might impact health professionals GBS knowledge | Almohaimeed et al., 2019; Gosling et al., 2002; Peralta-Carcelen et al. 1997; Price et al., 2018 | 4 | Gosling et al. (2002)  Peralta-Carcelen et al. (1997) | Almohaimeed et al. (2019) | Price et al. (2018) |  | **High confidence -**  most studies have no methodological concerns |
| 1.2.3 GBS as a public health issue  Most health professionals see GBS as an important public health issue. | The importance of GBS as a public health issue from health professional’s perspectives | Gosling et al., 2002; Price et al., 2018 | 2 | Gigante et al. (1995)  Gosling et al. (2002) | Almari et al. (2021)  Almohaimeed et al. (2019) | Price et al. (2018) |  | **Moderate confidence –** most of the studies had no or minor methodological concerns |
| 1.2.4 Screening as beneficial  Most health professionals see GBS screening as important and beneficial to pregnant women | The benefits of screening women for GBS | Alamri et al., 2021; Almohaimeed et al., 2019; Gosling et al., 2002; Price et al., 2018 | 4 | Gosling et al. (2002) | Almari et al. (2021)  Almohaimeed et al. (2019) | Price et al. (2018) |  | **Moderate confidence –** most of the studies had no or minor methodological concerns |
| **Review category: Preferences** | | | | | | | | |
| **2.1 Women’s views** | | | | | | | | |
| 2.1.1 Preference of GBS screening strategy  Most women surveyed are in favour of universal screening | Whether women prefer universal or risk-based screening | Alshengeti et al., 2020; Chow et al., 2013; Kolkman et al., 2017; Peralta-Cercelen et al., 1997 | 4 | Alshegenti et al. (2020)  Chow et al. (2013)  Constantinou et al. (2023)  Peralta-Carcelen et al. (1997) | Kolkman et al. (2017)  Sharpe et al. (2015) |  | De Mello et al. (2015) | **High confidence –** most of the studies had no methodological concerns |
| 2.1.2 Views in favour of screening  Most women surveyed would accept GBS testing and believe it is a good way to protect their baby | Women’s positive views about screening and testing for GBS | Chow et al., 2013; Constantinou et al., 2023; Kolkman et al., 2017; Sharpe et al., 2015 | 4 | Chow et al., 2013; Constantinou et al., 2023; | Kolkman et al., 2017; Sharpe et al., 2015 |  |  | **Moderate confidence –** equal split across confidence levels, so have gone for the middle |
| 2.1.3 Views against screening  Views against screening include embarrassment, fear of birth plans being altered, overmedicalization of birth and implications for their baby | Reasons women provide for not accepting GBS testing | Constantinou et al., 2023; De Mello et al., 2015; Kolkman et al., 2017; Sharpe et al., 2015 | 4 | Constantinou et al., 2023; | Kolkman et al., 2017;  Sharpe et al., 2015 |  | De Mello et al. (2015) | **Moderate confidence –** most of the studies had no or minor methodological concerns |
| 2.1.4 Views about swabbing  Over half of women surveyed would accept swabbing, and the provision of clear information is vital in mitigating anxiety | Women’s attitudes towards swabbing and testing positive | Chow et al., 2013; Constantinou et al., 2023; De Mello et al., 2015; Kolkman et al., 2017; Sharpe et al., 2015 | 5 | Chow et al., 2013; Constantinou et al., 2023; | Kolkman et al., 2017;  Sharpe et al., 2015 |  | De Mello et al. (2015) | **Moderate confidence –** most of the studies had no or minor methodological concerns |
| 2.1.5 Preferences for HCP vs self-swabbing  Preference for self-swabbing vs health professional swabbing varies across studies and countries. | The percentage of women surveys choosing health professional swabbing, self-swabbing, or no preference | Arya et al., 2008; Chen et al., 2020; Ka Ye Ko et al., 2016; Kolkman et al., 2017; Molnar et al., 1997; Price et al., 2006; Taylor et al., 1997; Torok and Dunn, 2000 | 7 |  | Chen et al. (2020)  Ka Ye Ko et al. (2016)  Kolkman et al. (2017)  Taylor et al. (1997)  Torok and Dunn (2000) | Molnar et al. (1997)  Price et al. (2006) | Arya et al. (2008) | **Moderate confidence –** most of the studies had minor methodological concerns |
| 2.1.6 Reasons for preferring self-swabbing  Reasons for self-swabbing include feeling in control, being more private, and feeling more physically comfortable. | Reasons surveyed women give for preferring self-swabbing | Chen et al., 2020; Sharpe et al., 2015; Taylor et al., 1997 | 3 |  | Chen et al. (2020)  Sharpe et al., 2015  Taylor et al. (1997) |  |  | **Moderate confidence –** most of the studies had minor methodological concerns |
| 2.1.7 Ease of self-swabbing  Women generally find self-swabbing easy and comfortable. | How easy women found self-swabbing | Ka Ye Ko et al., 2016; Mercer et al., 1995; Nebreda-Martin et al., 2022; Taylor et al., 1997 | 4 |  | Ka Ye Ko et al. (2016)  Nebreda-Martin et al. (2022)  Taylor et al. (1997) |  | Mercer et al. (1995) | **Moderate confidence –** most of the studies had minor methodological concerns |
| 2.1.8 Reasons for preferring clinician swabbing  If women prefer health professional swabbing, they do so because they are concerned about doing it wrong | Reasons women gave for preferring clinician swabbing | Arya et al., 2008; Ka Ye Ko et al., 2016; Kolkman et al., 2017; Nebreda-Martin et al., 2022; Price et al., 2006; Sharpe et al., 2015; Taylor et al., 1997; Torok and Dunn, 2000 | 8 |  | Ka Ye Ko et al. (2016)  Kolkman et al. (2017)  Nebreda-Martin et al. (2022)  Sharpe et al. (2015)  Taylor et al. (1997)  Torok and Dunn (2000) | Price et al. (2006) | Arya et al. (2008) | **Moderate confidence –** most of the studies had minor methodological concerns |
| 2.1.9 Demographics affecting preference  It is not clear what demographic characteristics impact swabbing preference | Demographic characteristics that might affect women’s swabbing preference | Chen et al., 2020; Molnar et al., 1997; Price et al., 2006 | 3 |  | Chen et al. (2020) | Molnar et al. (1997)  Price et al. (2006) |  | **Low confidence -** most have moderate methodological concerns |
| **2.2 Health professional’s attitudes** | | | | | | | | |
| 2.2.1 Preferred GBS strategy  It is not clear what screening method health professionals prefer. More research is needed. | Whether health professionals prefer universal or risk-based screening | Gigante et al., 1995; Gosling et al., 2002; Kolkman et al., 2017 | 3 | Gigante et al. (1995)  Gosling et al. (2002) | Kolkman et al. 2017 |  |  | **High confidence –** most of the studies had no methodological concerns |
| **Review Category: Acceptability** | | | | | | | | |
| **3.1 Women’s views** | | | | | | | | |
| 3.1.1 Acceptability of GBS screening  At least 80% of women find GBS swabbing acceptable | Percentages of women who feel screening and swabbing is acceptable. | Cheng et al., 2006; Chow et al., 2013; Daniels et al., 2010; Ka Ye Ko et al., 2016; Madrid et al., 2018 | 5 | Chow et al., 2013 | Cheng et al. (2006)  Ka Ye Ko et al. (2016) | Daniels et al. (2010) | Madrid et al. (2018) | **Moderate confidence –** most of the studies had minor methodological concerns |
| 3.1.2 Acceptability of vaginal vs rectal swabbing  Generally vaginal swabbing is more acceptable than anal swabbing | Percentage of women who find vaginal vs rectal swabbing acceptable | Chow et al., 2013; Daniels et al., 2010; Law et al., 2013 | 3 | Chow et al., 2013 | Law et al. (2013) | Daniels et al. (2010) |  | **Moderate confidence –** gone in the middle |
| 3.1.3 Women’s anxiety or worries surrounding GBS testing  Screening may increase anxiety in women, particularly the combined strategy. | Whether screening is associated with increased anxiety levels in women | Cheng et al., 2006; Daniels et al., 2009; Daniels et al., 2010; Kolkman et al., 2020 | 4 | Daniels et al. (2009) | Cheng et al. (2006) | Daniels et al. (2010)  Kolkman et al. (2020) |  | **Low confidence –** Most have moderate methodological concerns |
| 3.1.4 Facilitators to acceptability  Multiple demographics factors may influence GBS testing acceptability | Demographic and other factors that increase women’s views of acceptability of screening | Chow et al., 2013; Constantinou et al., 2023; Madrid et al., 2018 | 3 | Chow et al., 2013  Constantinou et al. 2023 |  |  | Madrid et al. (2018) | **Medium confidence –** have gone in the middle of high and very low |
| 3.1.5 Barriers to acceptability  Ethnicity and age may be associated with lower levels of acceptability | Demographic and other factors associated with women’s low levels of acceptability | Daniels et al., 2009; Daniels et al., 2010; Madrid et al., 2018; Sharpe et al., 2015 | 4 | Daniels et al. (2009) | Sharpe et al. (2015) | Daniels et al. (2010) | Madrid et al. (2018) | **Low confidence –** Have gone in the middle on the lower end |
| **3.2 Health professional’s views** | | | | | | | | |
| 3.2.1 Antenatal vs intrapartum screening  Intrapartum screening is potentially acceptable | Acceptability of rapid screening | Daniels et al., 2009; Mahieu et al., 2000; Melin et al., 2004 | 3 | Daniels et al. (2009) | Mahieu et al. (2000) | Melin et al. (2004) |  | **Moderate confidence –** have gone in the middle |
| 3.2.2 Universal vs risk based  It is not clear if health professionals find universal or risk-based screening more acceptable | Acceptability of universal screening vs risk based screening | Daniels et al., 2009; Kolkman et al., 2017; McLaughlin and Crowther, 2000; Melin et al., 2004; Yamaguchi and Ohashi, 2019 | 5 | Daniels et al. (2009) | Kolkman et al. (2017)  McLaughlin and Crowther (2000)  Yamaguchi and Ohashi (2019) | Melin et al. (2004) |  | **Moderate confidence –** most of the studies had minor methodological concerns |
| 3.3.3 Vaginal vs rectal swabs  Anal swabs are generally less acceptable than vaginal swabs | Health professionals’ views on the acceptability for the use of vaginal vs rectal swabs | Daniels et al., 2009; Mahieu et al., 2000 | 2 | Daniels et al. (2009) | Mahieu et al. (2000) |  |  | **Moderate confidence –** most of the studies had minor methodological concerns |
| 3.3.4 Acceptability of antibiotic treatment  Midwives appear to be opposed to universal antibiotic use, but obstetricians may be more for its use | Health professional’s views on the acceptability of universal antibiotic treatment | Daniels et al., 2009; Konrad et al., 2007 | 2 | Daniels et al. (2009) | Konrad et al. (2007) |  |  | **Moderate confidence –** most of the studies had minor methodological concerns |
| **Review category: Feasibility/Adherence** | | | | | | | | |
| **4.1 Women’s views** | | | | | | | | |
| 4.1.1 Adherence based on medical records  According to medical records, not all eligible women were swabbed, and 30.2-53% of swabs were caried out outside of recommended time points | The percentage of women who are swabbed for GBS based on medical records | Berikopoulou et al., 2021; De Mello et al., 2015; Jaworowski et al., 2016 | 3 | Berikopoulou et al. (2021) |  | Jaworowski et al. (2016) | De Mello et al. (2015) | **Low confidence –** most had moderate or serious concerns |
| 4.1.2 Women’s recollection of being offered or receiving GBS testing  Most women asked did not recall being offered or receiving GBS testing | Whether women remember the offer of GBS testing and whether they remember receiving it | Alamri et al., 2021; Berikopoulou et al., 2021; De Mello et al., 2015 | 3 | Berikopoulou et al. (2021) | Almari et al. (2021) |  | De Mello et al. (2015) | **Moderate confidence –** most studies had no or minor methodological concerns |
| 4.1.3 Medical records vs women’s recollection  Coherence between women’s recollection and medical records vary across studies | How often GBS testing information is recorded in women’s medical records compared to women’s recollection of testing | Berikopoulou et al., 2021; De Mello et al., 2015 | 2 | Berikopoulou et al. (2021) |  |  | De Mello et al. (2015) | **Low confidence –** have gone in the middle on the lower side |
| **4.2 Health professionals’ views** | | | | | | | | |
| 4.2.1 Barriers  Barriers to GBS screening programmes include organisational barriers, fear of the consequences (e.g., anxiety, overmedicalisation of birth); lack of clarity around guidelines; medicolegal reasons and lack of training. | Health professional’s views on barriers to screening and treating GBS | Alamri et al., 2021; Almohaimeed et al., 2019; Davies et al., 2001; Gosling et al., 2002; Kolkman et al., 2017; Price et al., 2018 | 6 | Gosling et al. (2002) | Almari et al. (2021)  Almohaimeed et al. (2019)  Davies et al. (2001)  Kolkman et al. (2017) | Price et al. (2018) |  | **Moderate confidence –** most of the studies had no or minor methodological concerns |
| 4.2.2. Facilitators  Facilitators to GBS screening programmes vary across studies | Factors health professionals believe are facilitators to screening and treating GBS | Davies et al., 2001; Konrad et al., 2007; Mahieu et al., 2000 | 3 |  | Davies et al. (2001)  Konrad et al. (2007)  Mahieu et al. (2000) |  |  | **Moderate confidence –** all studies had minor methodological concerns |
| 4.2.3 Adherence to screening protocols  Adherence to screening protocols varies across studies ranging from 21.3-100% for universal screening and 10-55% for screening under certain conditions | The percentages of health professionals or services that reported using a screening strategy | Alamri et al., 2021; Almohaimeed et al., 2019; Davies et al., 2001; Gosling et al., 2002; Konrad et al., 2007; Lynfield et al., 2000; Mahieu et al., 2000; Melin et al., 2004; Peralta-Carcelen et al., 1997 | 9 | Gosling et al. (2002)  Peralta-Carcelen et al. (1997) | Almari et al. (2021)  Almohaimeed et al. (2019)  Davies et al. (2001)  Konrad et al. (2007)  Mahieu et al. (2000) | Lynfield et al. (2000)  Melin et al. (2004) |  | **Moderate confidence –** most of the studies had no or minor methodological concerns |
| 4.2.4 Adherence to timing of screening protocols  There is not a clear pattern about whether health professionals adherence to vaginal vs rectal swabbing guidelines | When health professionals carry out swabbing | Davies et al., 2001; Lynfield et al., 2000; Yamaguchi and Ohashi, 2018 | 3 |  | Davies et al. (2001)  Yamaguchi and Ohashi (2019) | Lynfield et al. (2000) |  | **Moderate confidence –** most of the studies had minor methodological concerns |
| 4.2.5 Adherence to vaginal vs rectal swabbing  There is not a clear pattern about whether health professionals adherence to vaginal vs rectal swabbing guidelines | The number of health professionals who report vaginal vs rectal swabbing | Davies et al., 2001; Lynfield et al., 2000; | 2 |  | Davies et al. (2001) | Lynfield et al. (2000) |  | **Low confidence –** most studies had minor or moderate methodological concerns |
| 4.2.6 Adherence to antibiotic prophylaxis protocols  Positive test antibiotic use ranged from: 50-87%; positive screen + positive risk factor antibiotic use ranged from: 13-99%; Positive risk factor but no positive screen antibiotic use ranged from: 38-80% | The percentage of health professionals or service providers who reported using antibiotic treatment | Alamri et al., 2021; Almohaimeed et al., 2019; Davies et al., 2001; Konrad et al., 2007; Lynfield et al., 2000; Mahieu et al. 2000; McLaughlin & Crowther, 2000; Yamaguchi and Ohashi, 2018 | 8 |  | Almari et al. (2021)  Almohaimeed et al. (2019)  Davies et al. (2001)  Konrad et al. (2007)  Mahieu et al. (2000)  McLaughlin and Crowther (2000)  Yamaguchi and Ohashi (2019) | Lynfield et al. (2000) |  | **Moderate confidence –** most of the studies had minor methodological concerns |
| 4.2.7 Professional characteristics and adherence  Obstetricians and gynaecologists may be more likely to follow policies than nurses/midwives, and those who have worked less time may be more likely to follow policies | Characteristics that are associated with adherence to swabbing protocols and procedures | Alamri et al., 2021; Davies et al., 2001; Lynfield et al., 2000; Mahieu et al., 2000 | 4 |  | Alamri et al. (2021)  Davies et al. (2001)  Mahieu et al. (2000) | Lynfield et al. (2000) |  | **Moderate confidence –** most of the studies had minor methodological concerns |
| 4.2.8 Improving adherence  Most health professionals saw training as important for increasing adherence | Factors health professionals see as important for improving engagement with GBS protocols | Alamri et al., 2021; Almohaimeed et al., 2019; Price et al., 2006 | 3 |  | Alamri et al. (2021)  Almohaimeed et al. (2019)  Davies et al. (2001) |  |  | **Moderate confidence –** all studies had minor methodological concerns |
