## Supplemental Material E for "Acceptability and feasibility of maternal screening for Group B Streptococcus: a rapid review"

**GRADE CERQual Adequacy Table**

| **Statement of findings** | **Definition** | **Studies citing this** | **N** | **Summary of findings** | **Adequacy** | **CERQUAL Adequacy Rating** |
| --- | --- | --- | --- | --- | --- | --- |
| **Review category: Knowledge and awareness** | | | | | | |
| **1.1 Women’s views** | | | | | | |
| 1.1.1 Awareness   Awareness of GBS is generally low (<40%) and varies across countries and populations. | The percentage of women in the study sample with knowledge about GBS, GBS testing/screening and treatment for GBS. | Alamri et al., 2021; Alshengeti et al. 2020; Arya et al., 2008; Bak et al., 2016; Chow et al., 2013; Constantinou et al., 2023; Darbyshire et al., 2003; De Mello et al., 2015; Giles et al., 2019; Grammeniatis et al., 2022; Peralta-Carcelen et al., 1997; Sharpe et al., 2015; Youden et al., 2005 | 13 | *Awareness of GBS –*  **Alamri et al. (2021)** – 14.7% of the women from Saudi Arabia sampled had heard of GBS  **Arya et al. (2008)** – 25% of the women from Ireland sampled had heard of GBS  **Chow et al. (2013)** – 36% of the women from Hong Kong sampled had heard of GBS  **Constantinou et al. (2021)** – Qualitative data of 19 UK based women found that the sample had differing knowledge of GBS.  **Darbyshire et al. (2003)** – Qualitative data of 35 Australian based women found that a lot of the sample had no prior knowledge of GBS.  **Giles et al. (2019)** – 37% of the women from Australia sampled had heard of GBS  **Grammeniatis et al. (2022)** – 28% of the women from Greece sampled had heard of GBS  **Peralta-Carcelen et al. (1997)** – 8% of the women from the USA sampled had heard of GBS  **Sharpe et al. (2015)** – qualitative data of 6 Canadian women found the women had virtually no knowledge about GBS.  **Youden et al. (2005)** – women in the Canadian sample were significantly less knowledgeable about GBS infections than infections screened for during pregnancy, such as rubella, hepatitis B, and HIV.  *Knowledge about GBS testing –*  **Alamri et al. (2021)** – 9% of the women from Saudi Arabia sampled did not know that GBS testing was available during labour  **Bak et al., (2016)** – 67.1% of the women from Poland surveyed were aware that they had had a GBS test  **DeMello** **et al. (2015)** – 56.7% of the women from Brazil surveyed, were aware that they had had a GBS test  *Knowledge about treatment*  **Alshengeti et al. (2020)** – only 40% of the women from Saudi Arabia sampled knew when antibiotics should be given  **Chow et al. (2013)** – only 14% knew when antibiotics should be given | **Richness –** Most are statistics which is appropriate for this theme.  **Quantity -** 13 | **High adequacy –** a large amount of studies contributing to this theme |
| 1.1.1 Misconceptions about GBS   Misconceptions about GBS include believing it is an STD and it is women’s fault | Incorrect beliefs women may hold about GBS | Alamri et al., 2021; Alshengeti et al. 2020; Chow et al., 2013; Constantinou et al., 2023; Darbyshire et al., 2003; Grammeniatis et al., 2022; Sharpe et al., 2015; | 7 | *GBS as an STD*  **Alamri et al. (2021)** – 50.8% thought GBS was an STD  **Alshengeti et al. (2020)** – 60.5% thought GBS was an STD  **Chow et al. (2013)** – 13% thought that GBS was an STD  **Darbyshire et al. (2003)** – qualitative data, some women believed GBS was an STD  **Sharpe et al. (2015)** – qualitative data, some women believed GBS was an STD  *GBS as a leading cause of meningitis*  **Alamri et al. (2021)** – 40.8% thought it was a leading cause of meningitis  **Alshengeti et al. (2020)** – 62% thought it was a leading cause of meningitis  *Not understanding the seriousness of GBS*  **Darbyshire et al. (2003)** - qualitative data, most women were unclear about GBS, they tried to make sense of it through what illnesses it sounded like (strep throat) which made it difficult for them to gauge the severity of it.  **Grammeniatis et al. (2022)** – 55% did not know about the seriousness of GBS  *Women believing they are at fault for having GBS*  **Darbyshire et al. (2003)** - qualitative data, some women felt it was their fault for having GBS and wondered what they could have done differently  **Sharpe et al. (2015)** - qualitative data, some women felt it was their fault for having GBS and wondered what they could have done differently  *Other*  **Alamri et al. (2021)** – 28.3% of women thought GBS positive status meant women couldn’t breastfeed  **Chow et al. (2013)** – 72% overestimated the percentage of women in Hong Kong with GBS  **Constantinou et al. (2021)** – qualitative data, some women thought the swabbing was high vaginal swabbing and required the use of a speculum  **Sharpe et al. (2015)** – qualitative data, some women did not know they could decline IAP, and they were not aware that they had a choice over treatment | **Richness –** the qualitative papers provide more detail (which is appropriate).  **Quantity:** 7 | **Low adequacy -**  An appropriate amount of richness, but only 7 studies |
| 1.1.2 Where does women’s knowledge about GBS come from?   Women get information about GBS from a wide variety of sources (health professionals, books, journals, family and friends, social media) | Sources and locations that women access knowledge about GBS | Alamri et al., 2021; Constantinou et al., 2023; Darbyshire et al., 2003; De Mello et al. (2015); Sharpe et al., 2015; | 5 | **Alamri et al. (2021)** – Women got their information about GBS from doctors, social media, books, journals and researchers.  **Constantinou et al. (2021)** – qualitative data, women got their knowledge about GBS from their job, family/friends, antenatal education including online information, or personal experience.  **Darbyshire et al. (2003)** – qualitative data, women got their knowledge from information booklets but fuzzy about exact details, friends and family, or personal experience.  **DeMello** **et al. (2015)** - 74.4% of women got information about GBS screening from health professionals.  **Sharpe et al. (2015)** – qualitative data, women learnt about GBS through self-directed research and midwives. | **Richness:** An appropriate amount given the types of papers.  **Quantity:** 5 | **Low adequacy -** An appropriate amount of richness, but only 5 studies |
| 1.1.3 What knowledge do women want?   Women generally want detailed information about GBS delivered face to face, provided early enough to make informed decisions. | Information that women would like to receive about GBS and how they would like to receive this information | Alamri et al., 2021; Constantinou et al., 2023; De Mello et al. 2015; Kolkman et al. 2017; Sharpe et al., 2015; | 5 | **Constantinou et al. (2023) –** qualitative data, most women wanted more information about GBS and GBS testing. They would have preferred more detailed explanations, ideally given face to face. Some women reported that they only got information if they asked for it, and some health professionals played down a GBS positive status.  **Kolkman et al. (2017)** - 93% of their sample from The Netherlands wanted to receive general information about GBS in the first half of pregnancy, in the form of a leaflet or via a website, however they believed positive GBS status would be best delivered face-to-face.  *Poor delivery of information*  **Alamri et al. (2021)** - 86.6% of their sample were not informed about GBS testing from any health professional  **De Mello et al. (2015)** - 25.1% of women were not told about their result before labour, and 57.2% women reported they weren’t told what their GBS status meant for them.  **Sharpe et al. (2015)** - qualitative data, midwives would tell women GBS was normal, but this led to confusion when midwives then expressed the need for treatment. | **Richness:** Two studies provide a lot of detail.  **Quantity:** 5 | **Low adequacy -** An appropriate amount of richness, but only 5 studies |
| 1.1.5. Factors associated with knowledge  Higher levels of education appear to be associated with more knowledge about GBS. Other factors may impact knowledge | Demographic and health characteristics that might impact women’s GBS knowledge | Alamri et al., 2021; Alshengeti et al. 2020; Bak et al., 2016; Chow et al., 2013; Constantinou et al., 2023; Giles et al., 2019; Grammeniatis et al., 2022; Jaworowski et al., 2016; Youden et al., 2005 | 9 | *Age*  **Alamri et al. (2021)** – Younger women had more knowledge (not statistically significant)  **Alshengeti et al. (2020)** – Younger women had more knowledge  **Bak et al., (2016)** – The middle age group (26-30) had more knowledge about GBS testing, and the older age group (30+) had more knowledge about antibiotics and complications of GBS.  **Giles et al. (2019)** – Older women had more knowledge  **Youden et al. (2005)**  – Older women had more knowledge  *Education*  **Alamri et al. (2021)** – no relationship between knowledge and education  **Bak et al., (2016)** – higher level of education, the more knowledge women had  **Chow et al. (2013)** - higher level of education, the more knowledge women had  **Giles et al. (2019)** - higher level of education, the more knowledge women had  **Grammeniatis et al. (2022)** - higher level of education, the more knowledge women had  **Jaworowski et al. (2016)** – no relationship between education level and knowledge in the sample of women from Poland  **Youden et al. (2005)**  - higher level of education, the more knowledge women had  *Employment*  **Alamri et al. (2021)** – Employment was associated with lower levels of knowledge  **Chow et al. (2013)** – Those who were employed were more likely to have heard of GBS  **Giles et al. (2019)** – Those who were housewives had more knowledge about GBS  *Wealth*  **Chow et al. (2013)** – Those who had an income of higher than HK$20,000 had more knowledge  **Grammeniatis et al. (2022)** – Those with higher income had more knowledge  Jaworowski – Those with average or above wealth had more knowledge (but not statistically significant)  *Living area*  **Bak et al., (2016)** – Those living in the city were more likely to know about GBS testing  **Chow et al. (2013)** – Those who had lived in Hong Kong for longer than 7 years were more likely to have heard of GBS and have more knowledge about GBS  **Giles et al. (2019)** – Those born in Australia had more knowledge of GBS  **Jaworowski et al. (2016)** – There was no impact on whether women lived in a city or a village on knowledge about GBS  *Past exposure to GBS*  **Alshengeti et al. (2020)** – Women with past exposure to GBS had more knowledge  **Alamri et al. (2021)** – No relationship between past exposure to GBS and knowledge  **Constantinou et al. (2023)** – qualitative data, past exposure did not necessarily equate to more knowledge  **Jaworowski et al. (2016)** – women with past exposure to GBS had more knowledge  *Parity*  **Alamri et al. (2021)** – Parity and number of babies women had, had no relationship with knowledge  **Bak et al., (2016)** – Parity had no relationship with knowledge  **Jaworowski et al. (2016)** – Parity had no relationship with knowledge  **Alshengeti et al. (2020)** - multiparous women and women who had had more babies were more likely to be knowledgeable  **Giles et al. (2019)** – Women who had more than one child had more knowledge  *Other factors*  **Alshengeti et al. (2020)** – Women who had received antenatal care from a gynaecologist had more knowledge  **Jaworowski et al. (2016)** – Women who were married had more knowledge  **Chow et al. (2013)** – Women who had accepted GBS screening, and women who were in the third trimester of pregnancy (compared to the first) had more knowledge  **Youden et al. (2005) -** Women who had received culture based screening compared to risk based screening had more knowledge | **Richness –** Most are statistics which is appropriate for this theme.  **Quantity - 9** | **Moderate –** there are 9 studies in this theme and the richness is appropriate as all are quantitative. |
| **1.2 Health professionals** | | | | | | |
| 1.2.1 Awareness  Health professionals generally have higher knowledge about GBS, but some may be less aware of risk factors. | The percentage of health professionals the study sample with knowledge about GBS, GBS testing/screening and treatment for GBS. | Alamri et al., 2021; Almohaimeed et al., 2019; Gosling et al., 2002; Melin et al. 2004; Peralta-Carcelen et al., 1997; Price et al., 2018; | 6 | *Knowledge of screening*  **Alamri et al. (2021)** - 48% of physicians from the sample in Saudia Arabia, had good knowledge, and 28% had excellent knowledge of GBS screening. Only 56% of doctors answered correctly about only asymptomatic women being screened, and only 24% answered correctly about only patients with a history of GBS being screened.  **Almohaimeed et al. (2019)** - 31.5% of the sample from Saudi Arabia, had good knowledge and 57.3% had excellent knowledge of GBS screening. 41.6% of doctors answered correctly about only asymptomatic women being screened, and 66.3% answered correctly about only patients with a history of GBS being screened.  *Risk factors*  **Gosling et al., (2002)** - none of the health professionals asked (Midwives, GPs, obstetricians) from the sample in New Zealand were able to identify all five risk factors for GBS.  **Price et al., (2018)** - 77.7% of health professionals (obstetrics and nurses) from the sample in South Africa were unable to list a single risk factor for EOGBS.  *Management strategies*  **Melin et al. (2004)** - 83% of obstetricians knew what management strategy policy they used in their hospital in Belgium.  **Peralta-Carcelen et al. (1997)** - 80% of obstetricians in the USA were aware of the AAP strategy. 100% of obstetricians were aware of ACOG policy.  **Price et al. (2018)** - 22.7% of obstetricians and nurses from South Africa sampled identified the risk based screening approach, only 55% identified the correct antibiotic, and only 16.8% were aware of the appropriate time to give the antibiotic. | **Richness:** Alamri and Almohaimeed provide a lot of detail about this.  **Quantity:** 6 | **Low adequacy –** although two papers do provide good detail, there are only 6 papers in this theme |
| 1.2.2 Factors associated with knowledge  Obstetricians *may* have more knowledge about GBS than other health professionals (midwives, nurses, paediatricians). | Characteristics that might impact health professionals GBS knowledge | Almohaimeed et al., 2019; Gosling et al., 2002; Peralta-Carcelen et al. 1997; Price et al., 2018 | 4 | **Almohaimeed et al. (2019)** - Older obstetricians had more knowledge than younger ones.  **Gosling et al. (2002)** - midwives were more likely to say they were unaware of risk factors compared to GPs and obstetricians.  **Peralta-Carcelen et al. (1997)** - obstetricians had more knowledge than paediatricians about management strategies, this is despite more paediatricians managing a child with GBS, compared to percentage of obstetricians treating a GBS positive mother.  **Price et al. (2018)** - obstetricians were better at identifying risk factors and risk-based strategies than nurses. Nurses stated that they felt they did not have enough information about GBS or GBS prevention protocols and they would like to improve their knowledge via campaigns and media information. | **Richness:** Perala-Carcelen and Price provide a high level of detail  **Quantity:** 4 | **Very Low adequacy –** although two papers do provide good detail, there are only 4 papers in this theme |
| 1.2.3 GBS as a public health issue  Most health professionals see GBS as an important public health issue | The importance of GBS as a public health issue from health professional’s perspectives | Gosling et al., 2002; Price et al., 2018 | 2 | **Gosling et al. (2002)** - 79% of clinicians surveyed believed GBS infections are an important public health issue.  **Price et al. (2018)** – 100% of clinicians agreed or strongly agreed that GBS is an important public health issue. | **Richness:** there is an adequate amount considering quantitative papers  **Quantity:** 2 | **Very Low adequacy –** Only 2 papers |
| 1.2.4 Screening as beneficial  Most health professionals see GBS screening as important and beneficial to pregnant women | The benefits of screening women for GBS | Alamri et al., 2021; Almohaimeed et al., 2019; Gosling et al., 2002; Price et al., 2018 | 4 | **Alamri et al. (2021)** – 68% agreed or strongly agreed that it is important to screen for GBS. 72% agreed or strongly agreed that screening is beneficial for pregnant women. 82% would recommend screening.  **Almohaimeed et al. (2019) –** 91.1% agreed or strongly agreed that GBS screening is important. 88.8% agreed or strongly agreed that screening is beneficial for pregnant women, 82% would recommend screening.  **Gosling et al. (2002)** – 85% said screening for GBS is desirable  **Price et al. (2018)** – all clinicians agreed that GBS is an important cause of infection and the implementation of GBS protocol is important or very important. | **Richness:** Alamri and Almohaimeed have provided detail.  **Quantity:** 4 | **Very Low adequacy –** although two papers do provide good detail, there are only 4 papers in this theme |
| **Review category: Preferences** | | | | | | |
| **2.1 Women’s views** | | | | | | |
| 2.1.1 Preference of GBS screening strategy  Most women surveyed are in favour of universal screening | Whether women prefer universal or risk-based screening | Alshengeti et al., 2020; Chow et al., 2013; Kolkman et al., 2017; Peralta-Cercelen et al., 1997 | 4 | **Alshengeti et al. (2020) –** 61.8% of women believed that the universal culture based screening strategy was better than risk based  **Chow et al. (2013)** – 66% of women agreed with universal screening  **Kolkman et al. (2017) -**  86% of women preferred the combination strategy which is where all women are screened, but only those with a positive test and risk factor are prescribed antibiotics  **Peralta-Carcelen et al. (1997)** – 81% of the women preferred the AAP strategy (universal screenining) | **Richness:** Kolkman and Ashengeti provide a high level of detail  **Quantity:** 4 | **Very Low adequacy –** although two papers do provide good detail, there are only 4 papers in this theme |
| 2.1.2 Views in favour of GBS testing  Most women surveyed would accept GBS testing and believe it is a good way to protect their baby | Women’s positive views about screening and testing for GBS | Chow et al., 2013; Constantinou et al., 2023; Kolkman et al., 2017; Sharpe et al., 2015 | 4 | **Chow et al. (2013) -** 81% of women said they would be happy to accept GBS screening during their current pregnancy. Factors that were associated with women being more likely to accept testing were having lived in Hong Kong for over 7 years, being employed, being nulliparous and having previously heard of GBS.  **Constantinou et al. (2023) –** qualitative data, all but one of the women said they would accept swabbing, as long as there were no financial or physical costs to themselves or the baby. Most women believed that testing is beneficial because it was a good way to protect their baby. They also viewed the antibiotics as a simple way to prevent something from happening to their baby  **Kolkman et al. (2017) -** 57% of women would not be worried about a positive GBS status following testing because of the option of antibiotics to prevent adverse outcomes for their baby  **Sharpe et al. (2015) –** qualitative data which found that if information about safe treatment was provided, this could help with women’s anxiety following their positive test result. | **Richness:** Most papers provide a high level of detail  **Quantity:** 4 | **Very Low adequacy –** although there is richness in this theme, there are only 4 papers in this theme |
| 2.1.3 Views against testing  Views against screening include embarrassment, fear of birth plans being altered, overmedicalization of birth and implications for their baby | Reasons women provide for not accepting GBS testing | Constantinou et al., 2023; De Mello et al., 2015; Kolkman et al., 2017; Sharpe et al., 2015 | 4 | **Constantinou et al. (2023)** qualitative data, some women were against testing because it could lead to too much stress and anxiety being put on mothers, especially given the low rates of EOGBS. Women also wouldn’t accept because of the risk of over-medicalising birth and it impacting their birth plans, for example, the inability to give birth at home, or restricted movement due to the IV drip. Some women also stated they were concerned about the safety of swabbing, such as whether it would induce early labour, and the safety of antibiotics for their baby, especially during breastfeeding.  **De Mello et al. (2015)** - 7.3% of women felt embarrassed following swabbing, and 3% of women felt afraid.  **Kolkman et al. (2017)** - 29% of women would not participate in screening, because of the risk of over treatment (93%), and potential negative effects for the woman and her child (21%).  **Sharpe et al. (2015)** - qualitative data, some women responded with annoyance to their positive GBS test because they were concerned about the implications of it, such as needing to change their birth plan. | **Richness:** A high level of detail is provided by Constantinou  **Quantity:** 4 | **Very Low adequacy –** Low number of papers |
| 2.1.4 Views about swabbing  Over half of women surveyed would accept swabbing, and the provision of clear information is vital in mitigating anxiety | Women’s attitudes towards swabbing and testing positive | Chow et al., 2013; Constantinou et al., 2023; De Mello et al., 2015; Kolkman et al., 2017; Sharpe et al., 2015 | 5 | **Chow et al. (2013)** - 62% of women would accept low vaginal swabs. Women were less likely to accept high vaginal swabbing (only 30% would accept) and anal swabbing (only 13% would accept).  **Constantinou et al. (2023)** – qualitative data, most women saw the swabs as not particularly intrusive, but would want to be provided with high quality information about what it involved so they could make an informed choice. They wanted to know in depth information about what it would involve.  **De Mello et al. (2015) -** 68.3% felt indifferent after the test.  **Kolkman et al. (2017)** - 93% would accept swabbing.  **Sharpe et al. (2015) -** qualitative data, found that women believed the swabbing to be minimally invasive. Anxiety around the test could be mitigated with information, especially providing positive test results face-to-face. | **Richness:** De Mello and Kolkman have a low level of detail  **Quantity:** 5 | **Low adequacy –** although three papers do provide good detail, there are only 5 papers in this theme |
| 2.1.5 Preferences for HCP vs self-swabbing  Preference for self-swabbing vs health professional swabbing varies across studies and countries. | The percentage of women surveys choosing health professional swabbing, self-swabbing, or no preference | Arya et al., 2008; Chen et al., 2020; Ka Ye Ko et al., 2016; Kolkman et al., 2017; Molnar et al., 1997; Price et al., 2006; Taylor et al., 1997; Torok and Dunn, 2000 | 7 | **Arya et al. (2008)** – 43.2% of women had a strong preference for a health professional to collect their swab and would recommend health professional swabbing to a friend (45.2%)  **Chen et al. (2020)** – of the women surveyed in China, 20.9% preferred self swabbing, 53.6% had no preference, 25.3% preferred health professional swabbing  **Ka Ye Ko et al. (2016)** – of the women surveyed in China, 5.6% preferred self-swabbing, 31.9% had no preference, 62.5% preferred health professional  **Kolkman et al. (2017)** – 86% preferred clinician swabbing  Mercer et al. (1995) – of the women surveyed in the USA, 57.7% preferred self-swabbing  **Molnar et al. (1997)** – of the women surveyed in Canada, 34% preferred self swabbing, 41% had no preference, 26% preferred health professional  **Price et al. (2006)** – 27.3% preferred self swabbing, 56.3% had no preference, 22.7% preferred health professional  **Taylor et al. (1997)** – of the women surveyed in the USA, 57% preferred self swabbing, 6% had no preference, 37% preferred health professional swabbing  **Torok and Dunn (2000)** – of the women surveyed in the USA, 58% preferred self-swabbing | **Richness:** there is an adequate amount considering quantitative papers  **Quantity:** 7 | **Low adequacy –** richness is not high, but is fine as quantitative but only 7 studies |
| 2.1.6 Reasons for preferring self-swabbing  Reasons for self-swabbing include feeling in control, being more private, and feeling more physically comfortable. | Reasons surveyed women give for preferring self-swabbing | Chen et al., 2020; Sharpe et al., 2015; Taylor et al., 1997 | 3 | **Chen et al. (2020)** - fewer women reported pain during self sampling (11.9%) compared to health professional sampling (19.6%).  **Sharpe et al. (2015)** – qualitative data, the offer of self-swabbing meant women felt more in control of their care.  **Taylor et al. (1997)** - women who preferred self-swabbing did so because it offered them more privacy (57%) and greater physical comfort (10%). | **Richness:** there is an adequate amount considering quantitative papers  **Quantity:** 3 | **Very Low adequacy –** Only 3 papers |
| 2.1.7 Ease of self-swabbing  Women generally find self-swabbing easy and comfortable. | How easy women found self-swabbing | Ka Ye Ko et al., 2016; Mercer et al., 1995; Nebreda-Martin et al., 2022; Taylor et al., 1997 | 4 | **Ka Ye Ko et al. (2016)** -69.4% felt comfortable doing vaginal swab; 60.9% felt comfortable doing anal swab. Women who had previously used tampons before were more likely to feel they could complete vaginal self-swabbing, and having a preference for self-swabbing was also associated with women’s belief they could complete it.  **Mercer et al. (1995)** - 90.4% of the women from the USA sampled, did not find self-swabbing difficult, and 90% said they would like to be offered self-sampling in the future. Furthermore,  **Nebreda-Martin et al. (2022)** - 72.7% of the Spanish women found self-swabbing easy  **Taylor et al. (1997)** - 90% of women had no difficulty collecting their own swabs, with 77% describing the experience positively. | **Richness:**Ka Ye Ko provide a lot of detail.  **Quantity: 4** | **Very Low adequacy –** Only very rich paper, but only 4 studies |
| 2.1.8 Reasons for preferring clinician swabbing  If women prefer health professional swabbing, they do so because they are concerned about doing it wrong | Reasons women gave for preferring clinician swabbing | Arya et al., 2008; Ka Ye Ko et al., 2016; Kolkman et al., 2017; Nebreda-Martin et al., 2022; Price et al., 2006; Sharpe et al., 2015; Taylor et al., 1997; Torok and Dunn, 2000 | 8 | **Arya et al. (2008)** – The most common reason women gave for not wanting to do self-sampling was worrying they wouldn’t do it properly.  **Ka Ye Ko et al. (2016)** - Reasons given for preferring health professional swabbing: professionals have greater knowledge (60.5%), Worry about the accuracy of results (56.5%), Difficult to perform self-sampling (28.5%); Dislike the self-test (5.5%)  **Kolkman et al. (2017)** – women expected better outcomes and more physical difficulties if they did it themselves.  **Nebreda-Martin et al. (2022)** - reasons given for not wanting to do self-swabbing: I don't know if I'm doing it correctly (47%); Difficulty in introducing the swab into the two holes (31%) Difficulty with haemorrhoids(8,8%) I can't easily locate the holes(4.4%); I'm afraid I might injure my baby (2.9%) I find it unpleasant to manipulate my genitals (2.9%); Difficulty in handling the sample/instrumental (2.9%).  **Price et al. (2006)** -. Reasons giving for refusing the study: not liking the idea of self sampling (38.7%); 12/31), wanting only one test to be (22.6); feeling unable to do test (16.1).  **Sharpe et al. (2015) -** qualitative data, some women were unsure about how to perform the test, not sure if they are doing it properly and feeling shy about asking for more detailed information.  **Taylor et al. (1997)** - Thos who found it difficult gave the following reasons: 63% blamed the size of their bump; 38% had difficulty locating the anus.  **Torok and Dunn (2000)** - Of the patients who refused to participate, the most frequently cited reason was not wanting to bother with the extra time needed to perform repeated swab collection. The next most frequent reason was concern about collecting the swab accurately (anecdotal info) | **Richness –** Nebdreda-Martin provide a lot of detail. Ka Ye Ko, Sharpe and Torok and Dunn also provide some richness  **Quantity:** 8 | **Moderate adequacy –** some richness, but only 8 papers within this theme |
| 2.1.9 Demographics affecting preference  It is not clear what demographic characteristics impact swabbing preference | Demographic characteristics that might affect women’s swabbing preference | Chen et al., 2020; Molnar et al., 1997; Price et al., 2006 | 3 | **Chen et al. (2020)** - younger women preferred self sampling  **Molnar et al. (1997)** - younger women, women without medical training and those who had had a GBS swab done in a previous pregnancy were more likely to prefer that the physician take the swab.  **Price et al. (2006)** - women who had never been married, had less than high school education were more likely to refuse a study in which women would be requested to self-swab. | **Richness:** An appropriate amount of detail considering study design  **Quantity: 3** | **Very Low adequacy –** Only 3 studies |
| **2.2 Health professional’s views** | | | | | | |
| 2.2.1 Preferred GBS strategy  It is not clear what screening method health professionals prefer. More research is needed. | Whether health professionals prefer universal or risk-based screening | Gigante et al., 1995; Gosling et al., 2002; Kolkman et al., 2017 | 3 | **Gigante et al. (1995)** – 28% of health professionals surveyed in the USA believed in universal screening, because they did not see it as cost effective (39%); colonisation is not predictive of EOBS (31%) and treatment efficacy is not conclusive (24%). 95% believed in risk-based screening.  **Gosling et al. (2002)** - 61% preferred universal screening, 27% preferred risk-based and the remaining 12% were not sure, or believed that women should choose.  **Kolkman et al. (2017) –** 20 out of 27 care providers prefer the combination strategy (74%) | **Richness:** there is an adequate amount considering quantitative papers  **Quantity:** 3 | **Very Low adequacy –** Only 3 papers |
| **Review Category: Acceptability** | | | | | | |
| **3.1 Women’s views** | | | | | | |
| 3.1.1 Acceptability of GBS screening  At least 80% of women find GBS swabbing acceptable | Percentages of women who feel screening and swabbing is acceptable. | Cheng et al., 2006; Chow et al., 2013; Daniels et al., 2010; Ka Ye Ko et al., 2016; Madrid et al., 2018 | 5 | **Cheng et al. (2006) -** 100% said they felt positively about being screened for GBS in the current pregnancy and in any future pregnancies.  **Chow et al. (2013) –** 81% found GBS screening acceptable. 62% would be willing to pay for GBS testing  **Daniels et al. (2010) -** 80.5% satisfied or very satisfied with the information given; 94.3% happy or very happy with the way swabs were taken; 94.1% confident with its use in routine care  **Ka Ye Ko et al. (2016) -** 94.7% found an additional visit for GBS screening to be acceptable  **Madrid et al. (2018)-** Acceptability of collecting vaginal and rectovaginal swabs was 100% | **Richness:** an adequate amount of the study designs  **Quantity:** 5 | **Low adequacy:** there is an adequate amount of richness, but only 5 studies |
| 3.1.2 Acceptability of vaginal vs rectal swabbing  Generally vaginal swabbing is more acceptable than anal swabbing | Percentage of women who find vaginal vs rectal swabbing acceptable | Chow et al., 2013; Daniels et al., 2010; Law et al., 2013 | 3 | **Chow et al. (2013) –** 62% accepted low vaginal swabs. 30% accepted high vaginal swabs13% found anal swabbing acceptable  **Daniels et al. (2010) –** 82.4% found vaginal swabbing acceptable, and 70.1% found anal swabbing acceptable. 96.2% found vaginal swabbing less embarrassing.  **Law et al. (2013)–** 90% of women found lower vaginal swabbing acceptable. 84% found anal-rectal swabbing acceptable. | **Richness:** an adequate amount of the study designs  **Quantity:** 3 | **Very Low adequacy:** An adequate amount of richness but only 3 papers |
| 3.1.3 Women’s anxiety or worries surrounding GBS testing  Screening may increase anxiety in women, particularly the combined strategy. | Whether screening is associated with increased anxiety levels in women | Cheng et al., 2006; Daniels et al., 2009; Daniels et al., 2010; Kolkman et al., 2020 | 4 | **Cheng et al. (2006) –** GBS colonisation was associated with greater psychological distress on the state anxiety scores. This was despite no differences in anxiety pre-test. However, after birth, there were no differences in anxiety between groups.  **Daniels et al. (2009) –** there were no significant differences in stated anxiety between those who tested positive for GBS, those who were not tested, or those who tested negative.  **Daniels et al. (2010) –** there was no evidence that screening raised anxiety. The mean anxiety score was within the normal range.  **Kolkman et al. (2020) –** Cambridge Worry Scale scores were 0.82 before the implementation and 0.84 during the implementation (where they are given a leaflet about EOGBS and a GBS strategy was implemented). Worry scores were the highest where the combination strategy was implemented. GBS related worry scale scores was 0.90 before implementation and 0.90 during implementation. Women in the combination strategy region were more likely to score high on the probability of being colonised question and admission of baby to the neonatal ward question. Variables associated with highest worry scores during implementation were combination strategy, primiparity, non-Dutch origin, living alone, low SES. | **Richness:** There is a high level of detail provided by Kolkman et al.  **Quantity:** 4 | **Very Low adequacy:** Although there is richness of the data for one study, there are only 4 papers |
| 3.1.4 Facilitators to acceptability  Multiple demographics factors may influence GBS testing acceptability | Demographic and other factors that increase women’s views of acceptability of screening | Chow et al., 2013; Constantinou et al., 2023 Madrid et al., 2018 | 3 | **Chow et al. (2013)** - Participants were more likely to accept screening if they had resided in Hong Kong for ≥7 years, were employed, were nulliparous (p=0.020), and had heard about GBS (p=0.003)  **Constantinou et al. (2023) –** believing there are no financial or physical costs to them or the baby  **Madrid et al. (2018) -** Facilitators for acceptance included: a) Willingness to know whether they had a reproductive tract infection; b) Being interested in understanding the objectives of collecting vaginal and rectovaginalvagino-rectal samples; and c) Willingness to be treated and accompanied to the hospital in case of reproductive tract infection and avoiding transmitting them to their offsprings (Madrid) | **Richness:** both studies are detailed  **Quantity:** 2 | **Very Low adequacy:** Although there is richness of the data, there are only 3 papers |
| 3.1.5 Barriers to acceptability  Ethnicity and age may be associated with lower levels of acceptability | Demographic and other factors associated with women’s low levels of acceptability | Daniels et al., 2009; Daniels et al., 2010; Madrid et al., 2018; Sharpe et al., 2015 | 4 | **Daniels et al. (2009) –** Those with three or more pregnancies experienced less discomfort with vaginal swabs. British and Irish women reported greater levels of comfort with vaginal swabs than Indian or Pakistani women. White British and Irish groups and Black Caribbean women, were less embarrassed than Indian or Pakistani women. White British and Irish women were happier with sample taking than Indian or Pakistani women. Younger women found the test less acceptable, found vaginal and rectal swab more embarrassing, and found vaginal swabs less comfortable. Higher anxiety was associated with higher embarrassment. *Rapid testing:* White women and older women had more confidence the test would be carried out correctly, and that they would be able to make a free choice, than Indian, Pakistani and Black African women. White women were happier to be treated on the basis of the test thn Pakistani women. White women more likely to recommend the test. Risk status: medium risk were less embarrassed, high risk group were happier to have treatment on basis of the test. Higher salience meant less mebarassed, but also less confidence about the test being performed correctly and free to make choices.  **Daniels et al. (2010)** – South Asian and younger women were less likely to want to take part in the study about swabbing. White British, Irish and younger women were the most likely to decline because they objected to the swabs, particularly rectal ones.  **Madrid et al. (2018) -**  factors linked to low acceptability were fear of burning or pain, stigma, lack of privacy  **Sharpe et al. (2015) –** women felt unsure about screening because of fear it would affect birth plans, jeopardise home birth | **Richness:** There is a high level of detail provided by Daniels et al. (2009).  **Quantity:** 4 | **Very Low adequacy:** Although there is richness of the data for one study, there are only 4 papers |
| **3.2 Health professional’s views** | | | | | | |
| 3.2.1 Antenatal vs intrapartum screening  Intrapartum screening is potentially acceptable | Acceptability of rapid screening | Daniels et al., 2009; Mahieu et al., 2000; Melin et al., 2004 | 3 | **Daniels et al. 2009 –** qualitative, the midwives felt that taking samples during labour was fine. However, they would not let it interfere with womens care and would not take swabs if inappropriate. Practical issues with rapid testing in terms of multi-tasking or speed at which women labour. These issues were linked to staff shortages such as who would do the test. If it was part of routine practice there should be a dedicated person to do it. Some midwives thought rapid testing could be used alone, others thought it should be used in addition to risk based testing. Some midwives also said it may not be worth adding an addition task to be carried out during birth given the low levels of EOGBS.  **Mahieu et al. (2000) -** 4% did not believe in prenatal screening for GBS. 72% would screen at 35-37 weeks; 20% would screen at 26-28 weeks. 94% would use a simple culture test, 5% believed in rapid GBS testing.  **Melin et al. (2004) -** 47% voted for rapid screening when a clinically proven effective rapid test becomes available | **Richness:** There is a high level of detail provided by Daniels et al. (2009).  **Quantity:** 3 | **Very Low adequacy:** Although there is richness of the data for one study, there are only 3 papers |
| 3.2.2 Universal vs risk based  It is not clear if health professionals find universal or risk-based screening more acceptable | Acceptability of universal screening vs risk based screening | Daniels et al., 2009; Kolkman et al., 2017; McLaughlin and Crowther, 2000; Melin et al., 2004; Yamguchi et al., 2019 | 5 | **Daniels et al. 2009 –** qualitative data, most midwives were not sure about the risk based strategy because of the risk of overtreating people who do not need treatment, and then missing people who do need treatment.  **Kolkman et al., 2017–** 74% preferred the combination strategy. Universal antenatal screening least preferred. 89% would advice against its use because of negative side effects, over treatment, antibiotic resistance.  **Mclaughlin and Crowther (2000)–** 56% of obstetricians preferred universal screening; 61% of neonatologists preferred universal antenatal screening. Those who did not prefer said it wasn’t cost-effective, not always accurate predictor of neonatal disease.  **Melin et al. (2004) –** consensus meeting – 94% voted for a universal prenatal screening strategy of women at 35-37 weeks gestation.  **Yamaguchi and Ohashi (2019) –** 97.5% of paediatricians agreed with universal screening | **Richness:** Daniels and McLaughlin and Crowther provide detailed information  **Quantity:** 5 | **Low quality:** Although two studies provide richness, there are only 5 studies. |
| 3.3.3 Vaginal vs rectal swabs  Anal swabs are generally less acceptable than vaginal swabs | Health professionals views on the acceptability for the use of vaginal vs rectal swabs | Daniels et al., 2009; Mahieu et al., 2000 | 2 | **Daniels et al. (2009) –** qualitative data, all midwives deemed anal swabbing to be unacceptable, and they felt women felt similarly.  **Mahieu et al. (2000) -** Two studies looked at the acceptability of vaginal vs rectal swabs to health professionals and found that 61% of obstetricians would carry out vaginal swabbing, whereas only 27% would take swabs from both the vaginal and rectal area | **Richness:** An adequate level of detail is provided considering the study design  **Quantity: 2** | **Very Low adequacy:** Adequate detail, but only 2 studies |
| 3.3.4 Acceptability of antibiotic treatment  Midwives appear to be opposed to universal antibiotic use, but obstetricians may be more for its use | Health professional’s views on the acceptability of universal antibiotic treatment | Daniels et al., 2009; Konrad et al., 2017 | 2 | **Daniels et al. (2009)** - midwives were opposed to the universal administration of antibiotics to prevent GBS. They felt it was only appropriate if risk factor or tested positive. They felt pregnant women would be wary of antibiotic treatment because of resistance, and fears the medication will affect the baby. They were also concerned that intravenous antibiotics would interfere with normal birth plans.  **Konrad et al. (2017) –** when asked how many women they would prescribe antibiotics to prevent one neonatal GBS related death, 24% of obstetrician and 30% of family physicians said 10,000. 11% of obstetricians and 9% of family physicians said >50,000 | **Richness:** There is a high level of detail provided by Daniels et al. (2009).  **Quantity: 2** | **Very Low adequacy:** Although there is richness of the data for one study, there are only 2 papers |
| **Review category: Feasibility/Adherence** | | | | | | |
| **4.1 Women’s views** | | | | | | |
| 4.1.1 Adherence based on medical records  According to medical records, not all eligible women were swabbed, and 30.2-53% of swabs were caried out outside of recommended time points | The percentage of women who are swabbed for GBS based on medical records | Berikopoulou et al., 2021; De Mello et al., 2015; Jaworowski et al., 2016 | 3 | **Berikopoulou et al. (2021)** – 65.4% of women in Greece surveyed were swabbed for GBS. Reasons for not swabbing was because their obstetrician did not offer it to them.  **De Mello et al. (2015)** - only 49.8% of women had swabbing recorded on their medical card  **Jaworowski et al. (2016)** - 86% of women surveyed were swabbed for GBS. Reasons for not swabbing included ignorance and giving birth before the planned GBS test. | **Richness:** acceptable given the study designs  **Quantity:** 3 | **Very Low adequacy:** Adequate detail, but only 3 studies |
| 4.1.2 Women’s recollection of being offered or receiving GBS testing  Most women asked did not recall being offered or receiving GBS testing | Whether women remember the offer of GBS testing and whether they remember receiving it | Alamri et al., 2021; Berikopoulou et al., 2021; De Mello et al., 2015 | 3 | **Alamri et al. (2021)** - 79.1% of women were unaware of GBS screening assessments.  **Berikopoulou et al. (2021)** - 23% of women could not recall having a GBS test taken and 16.9% were unsure whether they did or not so did not answer the question.  **De Mello et al. (2015)** - 69.3% of women reported that they remembered being screened for GBS. | **Richness:** acceptable given the study designs  **Quantity:** 3 | **Very Low adequacy:** Adequate detail, but only 2 studies |
| 4.1.3 Medical records vs women’s recollection  Coherence between women’s recollection and medical records vary across studies | How often GBS testing information is recorded in women’s medical records compared to women’s recollection of testing | Berikopoulou et al., 2021; De Mello et al., 2015 | 2 | **Berikopoulou et al. (2021)** – medical records indicated 34.6% of women were not screened, and approximately 39.9% of women did not recall being screened or could not remember  **De Mello et al. (2015)** - 69.3% of women said they were screened, but only 49.8% had it recorded on their medical card. | **Richness:** acceptable given the study designs  **Quantity:** 2 | **Very Low adequacy:** Adequate detail, but only 2 studies |
| **4.2. Health professional’s views** | | | | | | |
| 4.2.1 Barriers  Barriers to GBS screening programmes include organisational barriers, fear of the consequences (e.g., anxiety, overmedicalisation of birth); lack of clarity around guidelines; medicolegal reasons and lack of training. | Health professional’s views on barriers to screening and treating GBS | Alamri et al., 2021; Almohaimeed et al., 2019; Davies et al., 2001; Gosling et al., 2002; Kolkman et al., 2017; Price et al., 2018 | 6 | **Alamri et al. (2021)** – barriers to screening, in order of most agreed with: systems and protocols, lack of training, lack of tools, fear of consequences  **Almohaimeed et al. (2019)** – The barriers for GBS screening are system and protocol (52.8%), lack of training (46.1%), lack of tools (24.7%), fear of consequences (14.6%), and other barriers (16.9%).  **Davies et al. (2001)** - The most common reasons given for not screening in 1997: were use of risk factors alone to guide prophylaxis (87%), and beliefs that there were no clear guideline (24%), that screening was not cost-effective (31%) and that screening was not useful (35%). Clinicians didn’t want to adopt screening earlier on in pregnancy because of medico-legal reasons and queries around effectiveness.  **Gosling et al. (2002)**– barriers to routine antenatal screening included: counselling and time constraints (11%), antibiotic resistance and creating anxiety (10%)  **Kolkman et al. (2017)** – determinants were reported for the combination strategy (n = 53) and the screening strategy (n = 49), followed by the risk-based strategy (n = 25) and the Dutch guideline (n = 21). The determinants were 3.6 times more often (116 times) reported as impeding than they were reported as facilitating (32 times). Most determinants were associated with the user (n = 71), followed by the guideline itself (n = 39), the organisational context (n = 34) and the socio-political context (n = 4).determinants that related to identifying risk factors were: procedural clarity, correctness of the guidelines and social support by other care providers. some participants were not fully aware about the content of the Dutch guideline and many recommendations in the current guideline lacked procedural clarity. Determinants related to screening for GBS colonisation were mainly related to the guideline itself (n = 6); the care provider (n = 14); and the organisational context (n = 7). Determinants related to swabbing during pregnancy were to do with it being time consuming because of anticipated anxiety by women, some women might dislike it because of discomfort. In terms of treatment, routine antibiotics in case of carrier status was expected to result in overtreatment, negative side effects such as an increase in antibiotic resistance, medicalization of birth and a decrease of the choices of women to opt for home birth.  **Price et al. (2018)** – supervisors did not appear to prioritise the GBS protocol | **Richness:** Kolkman et al provide particularly detailed and rich information  **Quantity:** 6 | **Low adequacy:** There is one paper that has very rich descriptions. However, there are only 6 papers. |
| 4.2.2. Facilitators  Facilitators to GBS screening programmes vary across studies | Factors health professionals believe are facilitators to screening and treating GBS | Davies et al., 2001; Konrad et al., 2017; Mahieu et al., 2000 | 3 | **Davies et al. (2001)**- 30% of physicians reported that they gave antibiotics antenatally to some or all women with GBS colonization. Reasons given were: GBS colonization, patient request or GBS disease in a previous neonate.  **Konrad et al. (2007)** – factors affected the choice of prevention strategy varied by health professional, but for obstetricians: literature (86%); community standard (67%); recommendation by society of obstetricians and gynaecologists of Canada (48%). Family physicians: community standards (65%); recommendations (61%); what peers and colleagues do (65%). Family medicine residents: literature (46%); community standard (42%); recommendations (71%); examples of instructors (75%).  **Mahieu et al. (2000) -** Many gynaecologists (24%) perform prenatal screening for a personal reason e.g. negative experience with GBS in the past, medicolegal reasons or at request of the parents and paediatricians. | **Richness:** There is a high level of detail provided by Konrad et al  **Quantity: 3** | **Very Low adequacy:** Although there is richness of the data for one study, there are only 3 papers |
| 4.2.3 Adherence to screening protocols  Adherence to screening protocols varies across studies ranging from 21.3-100% for universal screening and 10-55% for screening under certain conditions | The percentages of health professionals or services that reported using a screening strategy | Alamri et al., 2021; Almohaimeed et al., 2019; Davies et al., 2001; Gosling et al., 2002; Konrad et al., 2007; Lynfield et al., 2000; Mahieu et al., 2000; Melin et al., 2004; Peralta-Carcelen et al., 1997 | 9 | **Alamri et al. (2021)–** based on universal screening recommendations, 72% said they screened for GBS, 28% said they didn’t screen and 0% said they didn’t know  **Almohaimeed et al. (2019) -** based on universal screening recommendations, 21.3% said they did screening in the clinic, 66.3% said they didn’t, and the remaining 12.4% said they didn’t know  **Davies et al. (2001)–** in 1997 85% of clinicians were screening at least 75% of women  **Gosling et al. (2002) –** 71% said they did universal screening, and 23% said they did risk based screening  **Konrad et al. (2007) –** 100% of obstetricians did universal screening, only 66% of family physicians did universal screening  **Lynfield et al. (2000) –** 72% of clinicians in Connecticut used universal screening, 55% providers in Minnesota chose risk-based  **Mahieu et al. (2000) –** 44% believed in routine prenatal screening; 52% believed in screening in certain situations  **Melin et al. (2004) –** 78% had universal screening policy, 10% had mixed behaviour policy  **Peralta-Carcelen et al. (1997) –** 18% routinely screened all pregnant women, and 45% screened pregnant women under certain circumstances | **Richness:** acceptable given the study designs  **Quantity:** 9 | **Moderate adequacy:** There is an adequate level of detail considering study design, and 9 papers in this theme |
| 4.2.4 Adherence to timing of screening protocols  There is not a clear pattern about whether health professionals adherence to vaginal vs rectal swabbing guidelines | When health professionals carry out swabbing | Davies et al., 2001; Lynfield et al., 2000; Yamaguchi and Ohashi, 2018 | 3 | **Davies et al. (2001)-** a lot of physicians felt uncomfortable adopting a strategy to screen at 26-28 weeks gestational age. This was to do with medico-legal concerns about introducing a a vaginal examination early in the pregnancy and lack of evidence for effectiveness  **Lynfield -et al. (2000)** - 80-82% of cultures were collected with a week of the 35-37 weeks period.  **Yamaguchi -and Ohashi (2018)** - 47.5% of maternity professionals in maternity homes conducted the GBS screening test during 33–37 weeks following the JSOG guideline | **Richness:** acceptable given the study designs, Davies provides more detail  **Quantity:** 3 | **Very Low adequacy:** Adequate detail, but only 3 studies |
| 4.2.5 Adherence to vaginal vs rectal swabbing  There is not a clear pattern about whether health professionals adherence to vaginal vs rectal swabbing guidelines | The number of health professionals who report vaginal vs rectal swabbing | Davies et al., 2001; Lynfield et al., 2000; | 2 | **Davies et al. (2001)** – in 1994, 61% who screened antenatally used vaginal swabs, 25% used a combination of vaginal and rectal swabs. In 1997 61% used vaginal and rectal swabs  **Lynfield et al. (2000)** – 71% collected from both the vagina and rectum. Obstetricians and gynaecologists were more likely to collect from both compared to nurse midwives and family physicians. | **Richness:** acceptable given the study designs  **Quantity:** 2 | **Very Low adequacy:** Adequate detail, but only 2 studies |
| 4.2.6 Adherence to antibiotic prophylaxis protocols  Positive test antibiotic use ranged from: 50-87%; positive screen + positive risk factor antibiotic use ranged from: 13-99%; Positive risk factor but no positive screen antibiotic use ranged from: 38-80% | The percentage of health professionals or service providers who reported using antibiotic treatment | Alamri et al., 2021; Almohaimeed et al., 2019; Davies et al., 2001; Konrad et al., 2007; Lynfield et al., 2000; Mahieu et al. 2000; McLaughlin & Crowther, 2000; Yamaguchi and Ohashi, 2018 | 8 | **Alamri et al. (2021) –** 52% would give antibiotics in case of positive test  **Almomaiheed et al. (2019) -** 50% would give antibiotics in case of positive test  **Davies et al. (2001)**- 30% of clinicians would offer antenatal treatment, and 38% would start IAP if there was no positive result but risk factors were present  **Konrad et al. (2007)-**  87% obstetricians and 66% family physicians prescribed antibiotics in the case of a positive test. 13% of professionals only prescribed IAP to those GBS positive women if they also had other risk factors present.  **Lynfield et al. (2000) -** 80% said they would administer IAP for all five of the high-risk criteria (i.e., previous infant with invasive GBS disease, GBS bacteriuria during the current pregnancy, delivery at <37 weeks' gestation, duration of rupture of mem branes >18 hours, and intrapartum fever >100.4 F [>38 C])  **Mahieu et al. (2000)** - 39% would offer antenatal treatment, 32% would not treat, and 29% would treat if there were other risk factors. The risk factors influencing treatment with antibiotics were fever (57%), preterm or premature rupture of membranes (50%), or they were aware of a previous history of neonatal GBS disease (45%)  **McLaughlin and Crowther (2000) -** 97-99% of professionals would provide antibiotics if the woman had any of the following: preterm labour, premature rupture of membranes, or infection  **Yamaguchi and Ohashi (2018)** – 82.2% of midwives reported using IAP | **Richness:** acceptable given the study designs. Lynfield and Mahieu provide more detail  **Quantity:** 8 | **Moderate adequacy:** There is an adequate level of detail considering study design, and 8 papers in this theme |
| 4.2.7 Professional characteristics and adherence  Obstetricians and gynaecologists may be more likely to follow policies than nurses/midwives, and those who have worked less time may be more likely to follow policies | Characteristics that are associated with adherence to swabbing protocols and procedures | Alamri et al., 2021; Davies et al., 2001; Lynfield et al., 2000; Mahieu et al., 2000 | 4 | *Location*  **Lynfield et al. (2000)** – those in Connecticut more likely to use universal screening strategy than those in Minnesota. Those in Alberta were more likely to use a universal screening strategy than those in Toronto.  **Mahieu et al. (2000)** – Those living in West and East Flanders were least likely to agree with CDC universal screening recommendations compared to those living in Antwerp and Brabant in Belgium  *Time*  **Davies et al. (2001)**– universal screening increased from 1994 (79%) to 1997 (89%).  *Role*  **Alamri et al. (2021)**– 11.9% reported they would discuss GBS risks with women compared to 1.5% of nurses.  **Davies et al. (2001)** – obstetricians were less likely to report routine antenatal screening than obstetricians (in 1994, 80% v. 66%, this result was also evidenced in 1997, 87% v. 78% p < 0.001)  **Lynfield et al. (2000)** – Obstetricians and gynaecologists – more likely than family physician, nurse midwives to: report that they follow the five indicators of risk; state that their practice had a GBS policy in place, and follow the guidelines  *Time in practice*  **Davies et al. (2001)** – those who had been attending births for 15 years or less were more likely to practice universal screening  **Mahieu et al. (2000)** – older participants were less likely to agree with the CDC guidelines of universal screening. | **Richness:** Lynfield provide a good amount of detail.  **Quantity:** 4 | **Very Low adequacy:** Adequate detail, but only 4 studies |
| 4.2.8 Improving adherence  Most health professionals saw training as important for increasing adherence | Factors health professionals see as important for improving engagement with GBS protocols | Alamri et al., 2021; Almohaimeed et al., 2019; Price et al., 2006 | 3 | **Alamri et al. (2021)** – 40% of professionals were not trained, but 50% believed it to be an important training need  **Almohaimeed et al. (2019)** – 73% thought GBS training was an important need. Despite this, 64% said they did not train junior staff on this practice.  **Price et al (2006)** –health professionals deemed inclusion of GBS in their continuing education program as important. The clinicians also stated that engagement with the GBS protocols could be encouraged by receiving feedback regarding the wellbeing of a neonate that had been affected by GBS infection, as this could sensitise them to the issue. They also stated that campaigns and media information focused on GBS could be important in improving engagement | **Richness:** Price provides high level of detail  **Quantity: 3** | **Very Low adequacy:** Adequate detail, but only 3 studies |
