## Supplemental Material F for "Acceptability and feasibility of maternal screening for Group B Streptococcus: a rapid review"

**CERQUAL Relevance table**

| **Statement of findings** | **Definition** | **Studies citing this** | **N** | **High relevance**  Studies published between 2016-present | **Moderate relevance**  Studies published between 2002-2015 | **Low relevance –** studies published between 1996-2001 | **Very low relevance –** studies published before 1996 | **Overall relevance rating** |
| --- | --- | --- | --- | --- | --- | --- | --- | --- |
| **Review Category: Knowledge and awareness** | | | | | | | | |
| **1.1 Women’s views** | | | | | | | | |
| 1.1.1 Awareness   Awareness of GBS is generally low (<40%) and varies across countries and populations. | The percentage of women in the study sample with knowledge about GBS, GBS testing/screening and treatment for GBS. | Alamri et al., 2021; Alshengeti et al. 2020; Arya et al., 2008; Bak et al., 2016; Chow et al., 2013; Constantinou et al., 2023; Darbyshire et al., 2003; De Mello et al., 2015; Giles et al., 2019; Grammeniatis et al., 2022; Peralta-Carcelen et al., 1997; Sharpe et al., 2015; Youden et al., 2005 | 13 | Alamri et al. (2021); Alshengeti et al. (2020);  Bak et al. (2016)  Constantinou et al. (2023)  De Mello et al. (2015);  Giles et al. (2019); Grammeniatis et al. (2022); | Arya et al. (2008);  Chow et al. (2013)  Sharpe et al. (2015)  Youden et al. (2005) | Darbyshire et al. (2003);  Peralta-Carcelen et al. (1997) |  | **High relevance –** the majority of studies were published after WHO’s guidelines |
| 1.1.2 Misconceptions about GBS   Misconceptions about GBS include believing it is an STD and it is women’s fault | Incorrect beliefs women may hold about GBS | Alamri et al., 2021; Alshengeti et al. 2020; Chow et al., 2013; Constantinou et al., 2023; Darbyshire et al., 2003; Grammeniatis et al., 2022; Sharpe et al., 2015; | 7 | Alamri et al. (2021); Alshengeti et al. (2020);  Constantinou et al. (2023) | Chow et al. (2013)  Sharpe et al. (2015) | Darbyshire et al. (2003) |  | **Moderate relevance:** three published after, three published before WHO guidelines |
| 1.1.3 Where does women’s knowledge about GBS come from?   Women get information about GBS from a wide variety of sources (health professionals, books, journals, family and friends, social media) | Sources and locations that women access knowledge about GBS | Alamri et al., 2021; Constantinou et al., 2023; Darbyshire et al., 2003; De Mello et al. (2015); Sharpe et al., 2015; | 5 | Alamri et al. (2021)  Constantinou et al. (2023)  De Mello et al. (2015) | Sharpe et al. (2015) |  |  | **High relevance –** majority of the studies were published after WHO’s guidelines |
| 1.1.4 What knowledge do women want?   Women generally want detailed information about GBS delivered face to face, provided early enough to make informed decisions. | Information that women would like to receive about GBS and how they would like to receive this information | Alamri et al., 2021; Constantinou et al., 2023; De Mello et al. 2015; Kolkman et al. 2017; Sharpe et al., 2015; | 5 | Alamri et al. (2021)  Constantinou et al. (2023)  De Mello et al. (2015)  Kolkman et al. (2017) | Sharpe et al. (2015) |  |  | **High relevance –** majority of the studies were published after WHO’s guidelines |
| 1.1.5. Factors associated with knowledge  Higher levels of education appear to be associated with more knowledge about GBS. Other factors may impact knowledge | Demographic and health characteristics that might impact women’s GBS knowledge | Alamri et al., 2021; Alshengeti et al. 2020; Bak et al., 2016; Chow et al., 2013; Constantinou et al., 2023; Giles et al., 2019; Grammeniatis et al., 2022; Jaworowski et al., 2016; Youden et al., 2005 | 9 | Alamri et al. (2021); Alshengeti et al. (2020);  Bak et al. (2016)  Constantinou et al. (2023)  Giles et al. (2019)  Grammientiatis et al. (2022)  Jaworowski et al. (2016) | Chow et al. (2013);  Youden et al. (2005) |  |  | **High relevance –** the majority of studies were published after WHO’s guidelines |
| **1.2 Health professional’s views** | | | | | | | | |
| 1.2.1 Awareness  Health professionals generally have higher knowledge about GBS, but some may be less aware of risk factors. | The percentage of health professionals the study sample with knowledge about GBS, GBS testing/screening and treatment for GBS. | Alamri et al., 2021; Almohaimeed et al., 2019; Gosling et al., 2002; Melin et al. 2004; Peralta-Carcelen et al., 1997; Price et al., 2018; | 6 | Alamri et al. (2021); Almohaimeed et al. (2019);  Price et al. (2018) | Gosling et al. (2002)  Melin et al. (2004) | Peralta-Carcelen et al., (1997) |  | **Moderate relevance –** Three were published after WHO guidelines, but three published before |
| 1.2.2 Factors associated with knowledge  Obstetricians *may* have more knowledge about GBS than other health professionals (midwives, nurses, paediatricians). | Characteristics that might impact health professionals GBS knowledge | Almohaimeed et al., 2019; Gosling et al., 2002; Peralta-Carcelen et al. 1997; Price et al., 2018 | 4 | Almohaimeed et al., (2019)  Price et al., (2018) | Gosling et al., (2002); | Peralta-Carcelen et al. (1997) |  | **Moderate relevance–** Two were published after WHO guidelines, but two were published before |
| 1.2.3 GBS as a public health issue  Most health professionals see GBS as an important public health issue | The importance of GBS as a public health issue from health professional’s perspectives | Gosling et al., 2002; Price et al., 2018 | 2 | Price et al. (2018) | Gosling et al. (2002) |  |  | **Moderate relevance –** one published after WHO’s guidelines, one before |
| 1.2.4 Screening as beneficial  Most health professionals see GBS screening as important and beneficial to pregnant women | The benefits of screening women for GBS | Alamri et al., 2021; Almohaimeed et al., 2019; Gosling et al., 2002; Price et al., 2018 | 4 | Almari et al. (2021)  Almohaimeed et al. (2019)  Price et al. (2018) | Gosling et al. (2002) |  |  | **High relevance –** the majority published after WHO’s guidelines |
| **Review category: Preferences** | | | | | | | | |
| **2.1 Women’s views** | | | | | | | | |
| 2.1.1 Preference of GBS screening strategy  Most women surveyed are in favour of universal screening | Whether women prefer universal or risk-based screening | Alshengeti et al., 2020; Chow et al., 2013; Kolkman et al., 2017; Peralta-Cercelen et al., 1997 | 4 | Alshengeti et al. (2020)  Kolkman et al. (2017) | Chow et al. (2013) | Peralta-Carcelen et al. (1997) |  | **Moderate relevance–** Two were published after WHO guidelines, but two were published before |
| 2.1.2 Views in favour of GBS testing  Most women surveyed would accept GBS testing and believe it is a good way to protect their baby | Women’s positive views about screening and testing for GBS | Chow et al., 2013; Constantinou et al., 2023; Kolkman et al., 2017; Sharpe et al., 2015 | 4 | Constantinou et al. (2023)  Kolkman et al. (2017) | Chow et al. (2013)  Sharpe et al. (2015) |  |  | **Moderate relevance –** two published before and two published after WHO guidelines |
| 2.1.3 Views against testing  Views against screening include embarrassment, fear of birth plans being altered, overmedicalization of birth and implications for their baby | Reasons women provide for not accepting GBS testing | Constantinou et al., 2023; De Mello et al., 2015; Kolkman et al., 2017; Sharpe et al., 2015 | 4 | Constantinou et al. (2023)  Kolkman et al. (2017)  De Mello et al. (2015) | Sharpe et al. (2015) |  |  | **High relevance–** majority published after WHO’s guidelines |
| 2.1.4 Views about swabbing  Over half of women surveyed would accept swabbing, and the provision of clear information is vital in mitigating anxiety | Women’s attitudes towards swabbing and testing positive | Chow et al., 2013; Constantinou et al., 2023; De Mello et al., 2015; Kolkman et al., 2017; Sharpe et al., 2015 | 5 | Constantinou et al., (2023)  De Mello et al. (2015)  Kolkman et al., (2017) | Chow et al., (2013);  Sharpe et al., (2015) |  |  | **High relevance –** the majority published after WHO’s guidelines |
| 2.1.5 Preferences for HCP vs self-swabbing  Preference for self-swabbing vs health professional swabbing varies across studies and countries. | The percentage of women surveys choosing health professional swabbing, self-swabbing, or no preference | Arya et al., 2008; Chen et al., 2020; Ka Ye Ko et al., 2016; Kolkman et al., 2017; Molnar et al., 1997; Price et al., 2006; Taylor et al., 1997; Torok and Dunn, 2000 | 7 | Chen et al. (2020)  Ka Ye Ko et al. (2016)  Kolkman et al. (2017) | Arya et al. (2008)  Price et al. (2006) | Molnar et al. (1997)  Taylor et al. (1997)  Torok and Dunn (2000) |  | **Moderate relevance –** three with high relevance, three with low relevance, have gone in the middle |
| 2.1.6 Reasons for preferring self-swabbing  Reasons for self-swabbing include feeling in control, being more private, and feeling more physically comfortable. | Reasons surveyed women give for preferring self-swabbing | Chen et al., 2020; Sharpe et al., 2015; Taylor et al., 1997 | 3 | Chen et al. (2020) | Sharpe et al., (2015) | Taylor et al. (1997) |  | **Moderate relevance–** one high, one moderate and one low so have chosen the middle |
| 2.1.7 Ease of self-swabbing  Women generally find self-swabbing easy and comfortable. | How easy women found self-swabbing | Ka Ye Ko et al., 2016; Mercer et al., 1995; Nebreda-Martin et al., 2022; Taylor et al., 1997 | 4 | Ka Ye Ko et al. (2016)  Nebreda-Martin et al. (2022) |  | Taylor et al. (1997) | Mercer et al. (1995) | **Moderate relevance –** two with high relevance, two with lower relevance so have gone in the middle. |
| 2.1.8 Reasons for preferring clinician swabbing  If women prefer health professional swabbing, they do so because they are concerned about doing it wrong | Reasons women gave for preferring clinician swabbing | Arya et al., 2008; Ka Ye Ko et al., 2016; Kolkman et al., 2017; Nebreda-Martin et al., 2022; Price et al., 2006; Sharpe et al., 2015; Taylor et al., 1997; Torok and Dunn, 2000 | 8 | Ka Ye Ko et al. (2016)  Kolkman et al. (2017)  Nebreda-Martin et al. (2022) | Sharpe et al. (2015)  Price et al. (2006)  Arya et al. (2008) | Taylor et al. (1997)  Torok and Dunn (2000) |  | **Moderate relevance –** three with high relevance, three with moderate and two with low so have gone in the middle |
| 2.1.9 Demographics affecting preference  It is not clear what demographic characteristics impact swabbing preference | Demographic characteristics that might affect women’s swabbing preference | Chen et al., 2020; Molnar et al., 1997; Price et al., 2006 | 3 | Chen et al. (2020) | Price et al. (2006) | Molnar et al. (1997) |  | **Moderate relevance–** one high, one moderate and one low so have chosen the middle |
| **2.2 Health professional’s attitudes** | | | | | | | | |
| 2.2.1 Preferred GBS strategy  It is not clear what screening method health professionals prefer. More research is needed. | Whether health professionals prefer universal or risk-based screening | Gigante et al., 1995; Gosling et al., 2002; Kolkman et al., 2017 | 3 | Kolkman et al., (2017) | Gosling et al. (2002) |  | Gigante et al. (1995) | **Low relevance –** Have gone in the middle |
| **Review Category: Acceptability** | | | | | | | | |
| **3.1 Women’s views** | | | | | | | | |
| 3.1.1 Acceptability of GBS screening  At least 80% of women find GBS swabbing acceptable | Percentages of women who feel screening and swabbing is acceptable. | Cheng et al., 2006; Chow et al., 2013; Daniels et al., 2010; Ka Ye Ko et al., 2016; Madrid et al., 2018 | 5 | Ka Ye Ko et al. (2016)  Madrid et al. (2018) | Cheng et al. (2006)  Chow et al., (2013)  Daniels et al. (2010) |  |  | **Moderate relevance –** most studies were published between 2002-2015 |
| 3.1.2 Acceptability of vaginal vs rectal swabbing  Generally vaginal swabbing is more acceptable than anal swabbing | Percentage of women who find vaginal vs rectal swabbing acceptable | Chow et al., 2013; Daniels et al., 2010; Law et al., 2013 | 3 |  | Chow et al., (2013)  Daniels et al. (2010)  Law et al. (2013) |  |  | **Moderate relevance –** all studies are of moderate relevance |
| 3.1.3 Women’s anxiety or worries surrounding GBS testing  Screening may increase anxiety in women, particularly the combined strategy. | Whether screening is associated with increased anxiety levels in women | Cheng et al., 2006; Daniels et al., 2009; Daniels et al., 2010; Kolkman et al., 2020 | 4 | Kolkman et al. (2020) | Cheng et al. (2006)  Daniels et al. (2009)  Daniels et al. (2010) |  |  | **Moderate relevance –** most studies were published between 2002-2015 |
| 3.1.4 Facilitators to acceptability  Multiple demographics factors may influence GBS testing acceptability | Demographic and other factors that increase women’s views of acceptability of screening | Chow et al., 2013; Constantinou et al., 2023; Madrid et al., 2018 | 2 | Madrid et al. (2018)  Constantinou et al. (2023) | Chow et al., (2013) |  |  | **High relevance -**  most in high relevance category |
| 3.1.5 Barriers to acceptability  Ethnicity and age may be associated with lower levels of acceptability | Demographic and other factors associated with women’s low levels of acceptability | Daniels et al., 2009; Daniels et al., 2010; Madrid et al., 2018; Sharpe et al., 2015 | 4 | Sharpe et al. (2015)  Madrid et al. (2018) | Daniels et al. (2009)  Daniels et al. (2010) |  |  | **Moderate relevance -**  two high, two moderate relevance |
| **3.2 Health professional’s views** | | | | | | | | |
| 3.2.1 Antenatal vs intrapartum screening  Intrapartum screening is potentially acceptable | Acceptability of rapid screening | Daniels et al., 2009; Mahieu et al., 2000; Melin et al., 2004 | 3 |  | Daniels et al. (2009)  Melin et al. (2004) | Mahieu et al. (2000) |  | **Moderate relevance –** most studies were published between 2002-2015 |
| 3.2.2 Universal vs risk based  It is not clear if health professionals find universal or risk-based screening more acceptable | Acceptability of universal screening vs risk based screening | Daniels et al., 2009; Kolkman et al., 2017; McLaughlin and Crowther, 2000; Melin et al., 2004; Yamaguchi and Ohashi, 2019 | 5 | Kolkman et al. (2017)  Yamaguchi and Ohashi (2019) | Daniels et al. (2009)  Melin et al. (2004) | McLaughlin and Crowther (2000) |  | **Moderate relevance –** two high, two moderate, one low so have gone in middle |
| 3.3.3 Vaginal vs rectal swabs  Anal swabs are generally less acceptable than vaginal swabs | Health professionals views on the acceptability for the use of vaginal vs rectal swabs | Daniels et al., 2009; Mahieu et al., 2000 | 2 |  | Daniels et al. (2009) | Mahieu et al. (2000) |  | **Low relevance –** one moderate, one low relevance |
| 3.3.4 Acceptability of antibiotic treatment  Midwives appear to be opposed to universal antibiotic use, but obstetricians may be more for its use | Health professional’s views on the acceptability of universal antibiotic treatment | Daniels et al., 2009; Konrad et al., 2007 | 2 |  | Daniels et al. (2009)  Konrad et al. (2007) |  |  | **Moderate relevance –** all published between 2002-2015 |
| **Review category: Feasibility/Adherence** | | | | | | | | |
| **4.1 Women’s views** | | | | | | | | |
| 4.1.1 Adherence based on medical records  According to medical records, not all eligible women were swabbed, and 30.2-53% of swabs were caried out outside of recommended time points | The percentage of women who are swabbed for GBS based on medical records | Berikopoulou et al., 2021;  De Mello et al. 2015; Jaworowski et al., 2016 | 3 | Berikopoulou et al. (2021)  Jaworowski et al. (2016) | De Mello et al. (2015) |  |  | **High relevance –** all studies published after WHO guidelines |
| 4.1.2 Women’s recollection of being offered or receiving GBS testing  Most women asked did not recall being offered or receiving GBS testing | Whether women remember the offer of GBS testing and whether they remember receiving it | Alamri et al., 2021; Berikopoulou et al., 2021; De Mello et al., 2015 | 3 | Almari et al. (2021)  Berikopoulou et al. (2021) | De Mello et al. (2015) |  |  | **High relevance –** most published after WHO guidelines |
| 4.1.3 Medical records vs women’s recollection  Coherence between women’s recollection and medical records vary across studies | How often GBS testing information is recorded in women’s medical records compared to women’s recollection of testing | Berikopoulou et al., 2021; De Mello et al., 2015 | 2 | Berikopoulou et al. (2021) | De Mello et al. (2015) |  |  | **Moderate relevance –** one high relevance, one moderate |
| **4.2 Health professional’s views** | | | | | | | | |
| 4.2.1 Barriers  Barriers to GBS screening programmes include organisational barriers, fear of the consequences (e.g., anxiety, overmedicalisation of birth); lack of clarity around guidelines; medicolegal reasons and lack of training. | Health professional’s views on barriers to screening and treating GBS | Alamri et al., 2021; Almohaimeed et al., 2019; Davies et al., 2001; Gosling et al., 2002; Kolkman et al., 2017; Price et al., 2018 | 6 | Almari et al. (2021)  Almohaimeed et al. (2019)  Kolkman et al. (2017)  Price et al. (2018) | Gosling et al. (2002) | Davies et al. (2001) |  | **High relevance –** most published after WHO guidelines |
| 4.2.2. Facilitators  Facilitators to GBS screening programmes vary across studies | Factors health professionals believe are facilitators to screening and treating GBS | Davies et al., 2001; Konrad et al., 2007; Mahieu et al., 2000 | 3 |  | Konrad et al. (2007) | Davies et al. (2001)  Mahieu et al. (2000) |  | **Low relevance –** most have low relevance |
| 4.2.3 Adherence to screening protocols  Adherence to screening protocols varies across studies ranging from 21.3-100% for universal screening and 10-55% for screening under certain conditions | The percentages of health professionals or services that reported using a screening strategy | Alamri et al., 2021; Almohaimeed et al., 2019; Davies et al., 2001; Gosling et al., 2002; Konrad et al., 2007; Lynfield et al., 2000; Mahieu et al., 2000; Melin et al., 2004; Peralta-Carcelen et al., 1997 | 9 | Almari et al. (2021)  Almohaimeed et al. (2019) | Gosling et al. (2002)  Konrad et al. (2007)  Melin et al. (2004) | Davies et al. (2001)  Lynfield et al. (2000)  Mahieu et al. (2000)  Peralta-Carcelen et al. (1997) |  | **Low relevance –** most studies published between 1996-2001 |
| 4.2.4 Adherence to timing of screening protocols  There is not a clear pattern about whether health professionals adherence to vaginal vs rectal swabbing guidelines | When health professionals carry out swabbing | Davies et al., 2001; Lynfield et al., 2000; Yamaguchi and Ohashi, 2018 | 3 | Yamaguchi and Ohashi (2019) |  | Davies et al. (2001)  Lynfield et al. (2000) |  | **Low relevance –** most studies published between 1996-2001 |
| 4.2.5 Adherence to vaginal vs rectal swabbing  There is not a clear pattern about whether health professionals adherence to vaginal vs rectal swabbing guidelines | The number of health professionals who report vaginal vs rectal swabbing | Davies et al., 2001; Lynfield et al., 2000; | 2 |  |  | Davies et al. (2001)  Lynfield et al. (2000) |  | **Low relevance –** all studies published between 1996-2001 |
| 4.2.6 Adherence to antibiotic prophylaxis protocols  Positive test antibiotic use ranged from: 50-87%; positive screen + positive risk factor antibiotic use ranged from: 13-99%; Positive risk factor but no positive screen antibiotic use ranged from: 38-80% | The percentage of health professionals or service providers who reported using antibiotic treatment | Alamri et al., 2021; Almohaimeed et al., 2019; Davies et al., 2001; Konrad et al., 2007; Lynfield et al., 2000; Mahieu et al. 2000; McLaughlin & Crowther, 2000; Yamaguchi and Ohashi, 2018 | 8 | Almari et al. (2021)  Almohaimeed et al. (2019)  Yamaguchi and Ohashi (2019) | Konrad et al. (2007) | Davies et al. (2001)  Lynfield et al. (2000)  Mahieu et al. (2000)  McLaughlin and Crowther (2000) |  | **Low relevance –** most studies published between 1996-2001 |
| 4.2.7 Professional characteristics and adherence  Obstetricians and gynaecologists may be more likely to follow policies than nurses/midwives, and those who have worked less time may be more likely to follow policies | Characteristics that are associated with adherence to swabbing protocols and procedures | Alamri et al., 2021; Davies et al., 2001; Lynfield et al., 2000; Mahieu et al., 2000 | 4 | Alamri et al. (2021) |  | Davies et al. (2001)  Lynfield et al. (2000)  Mahieu et al. (2000) |  | **Low relevance –** most studies published between 1996-2001 |
| 4.2.8 Improving adherence  Most health professionals saw training as important for increasing adherence | Factors health professionals see as important for improving engagement with GBS protocols | Alamri et al., 2021; Almohaimeed et al., 2019; Price et al., 2006 | 3 | Alamri et al. (2021)  Almohaimeed et al. (2019) |  | Davies et al. (2001) |  | **High relevance –** most published after WHO guidelines |
